# Genome-wide analysis of brain age identifies 25 associated loci and unveils relationships with mental and physical health

**DOI:** 10.1101/2023.12.26.23300533

**Authors:** Philippe Jawinski, Helena Forstbach, Holger Kirsten, Frauke Beyer, Arno Villringer, A. Veronica Witte, Markus Scholz, Stephan Ripke, Sebastian Markett

## Abstract

Neuroimaging and machine learning are opening up new opportunities in studying biological aging mechanisms. In this field, ‘brain age gap’ has emerged as promising MRI-based biomarker quantifying the deviation between an individual’s biological and chronological age of the brain – an indicator of accelerated/decelerated aging. Here, we investigated the genetic architecture of brain age gap and its relationships with over 1,000 health traits. Genome-wide analyses in 32,634 UK Biobank individuals unveiled a 30% SNP-based heritability and highlighted 25 associated loci. Of these, 23 showed sign-consistency and 16 replicated in another 7,259 individuals. The leading locus encompasses *MAPT*, encoding the tau protein central to Alzheimer’s disease. Genetic correlations revealed relationships with various mental health (depression), physical health (diabetes), and socioeconomic variables (education). Mendelian Randomization indicated a causal role of enhanced blood pressure on accelerated brain aging. This work refines our understanding of genetically modulated brain aging and its implications for human health.

## Main

Aging is an intricate biological phenomenon inherent to most living organisms.^1–3^ With extended human lifespans and the rapid pace of global demographic aging, age-related disabilities including neurodegenerative disorders such as dementia are on the rise.^4^ Understanding the biological mechanisms of aging is thus an urgent priority for health and social systems, to sustain longer lives with reduced periods of disability.

The use of neuroimaging methods in conjunction with machine learning has become a promising avenue in biomedical research to capture an individual’s biological age, with particular emphasis put on ‘brain age’.^5,6^ Brain age is typically assessed by training an age prediction model on in-vivo MRI data from a normative lifespan sample. This model is then applied to MRI data of unseen individuals to predict their age. The discrepancy between an individual’s brain-predicted and chronological age is termed ‘brain age gap’ (BAG), and is used to draw inferences on typical and atypical aging trajectories.^6,7^

A positive BAG (interpreted as accelerated aging) has been linked to reduced mental and physical fitness;^5^ including weaker grip strength, higher blood pressure, diabetes, adverse drinking and smoking behavior, poorer cognitive abilities, and depressive symptoms.^8–13^ Enhanced BAG is also evident in neurological and psychiatric disorders such as Alzheimer’s disease, schizophrenia, and bipolar disorder.^14,15^ While previous genetic studies suggest that BAG exhibits a substantial heritable component, only few studies have identified specific genetic variants that contribute to BAG.^15–21^ To refine the genetic architecture of BAG and identify potential therapeutic targets for healthy aging, further research is imperative.

In this report, we present what is to our knowledge the largest genome-wide association study (GWAS) of BAG to date. We discover novel loci in a sample of 32,634 individuals of white-British ancestry, and replicate our findings in a cross-ancestry sample of 7,259 individuals. This constitutes a 34% increase in sample size (about 10,000 more) compared to the most recent study^20^ First, we prioritize genes using complementary fine-mapping, annotation, and co-localization strategies that integrate information from multiple omics resources. Second, we replicate variant effects and calculate polygenic scores to estimate the present yield in variance explanation. Third, we compute genetic correlations with over 1,000 health traits. Fourth, we use Mendelian Randomization to test a potential causal role of several risk factors in BAG. Finally, we examine the degree of polygenicity and project discoveries to forthcoming studies. By these efforts, we unravel new biological mechanisms behind BAG, such as binding of small GTPases, i.e., evolutionary conserved proteins that act as biological timers of cellular processes.

## Results

Our brain age estimation workflow adhered to a well-established approach that utilizes the CAT12 voxel-based morphometry pipeline and has been validated extensively over the past years.^5,22^ We used T1-weighed anatomical MRI scans and machine learning in a cross-validation manner to estimate brain age in a discovery sample of 32,634 white-British ancestry individuals of the UK Biobank (UKB) cohort (age range: 45-82 years).^23^ Since brain aging has been demonstrated to encompass biologically distinct modes of change, we performed tissue-specific analyses on grey and white matter segmentations to increase the yield of biologically meaningful markers.^18^ Machine learning was carried out using complementary algorithms: the sparse Bayesian relevance vector machine,^24^ and the extreme gradient boosting technique (XGBoost) applied with both a tree and linear booster.^25^ Trained models were stacked within and across tissue classes to enhance prediction accuracy. This revealed three brain-predicted age estimates per subject, representing the age prediction for grey matter, white matter, and combined grey and white matter.

In the discovery sample, we observed excellent prediction accuracies of chronological age, with mean absolute errors (MAE) reaching MAE = 3.09 years and correlation coefficients attaining *r* = .86 (Fig. 1a, Table 1; details in suppl. Figure A1 and suppl. Table B1). Model performances were similar in two replication samples: a cross-ancestry UKB sample (*n* = 5,427; age range: 45-80 years), and a European ancestry sample drawn from the LIFE-Adult cohort (*n* = 1,833; age range: 45-80 years).^26,27^ Noteworthy, genetic association analyses were performed on brain age gap (BAG), i.e., the discrepancy between an individual’s brain-predicted and chronological age. These estimates – regressed on sex, age, age^2^, scanner site, and total intracranial volume – showed excellent test-retest reliabilities, with intra class correlation coefficients (ICC C,1)^28^ ranging from .89 to .92.

**Fig. 1.**
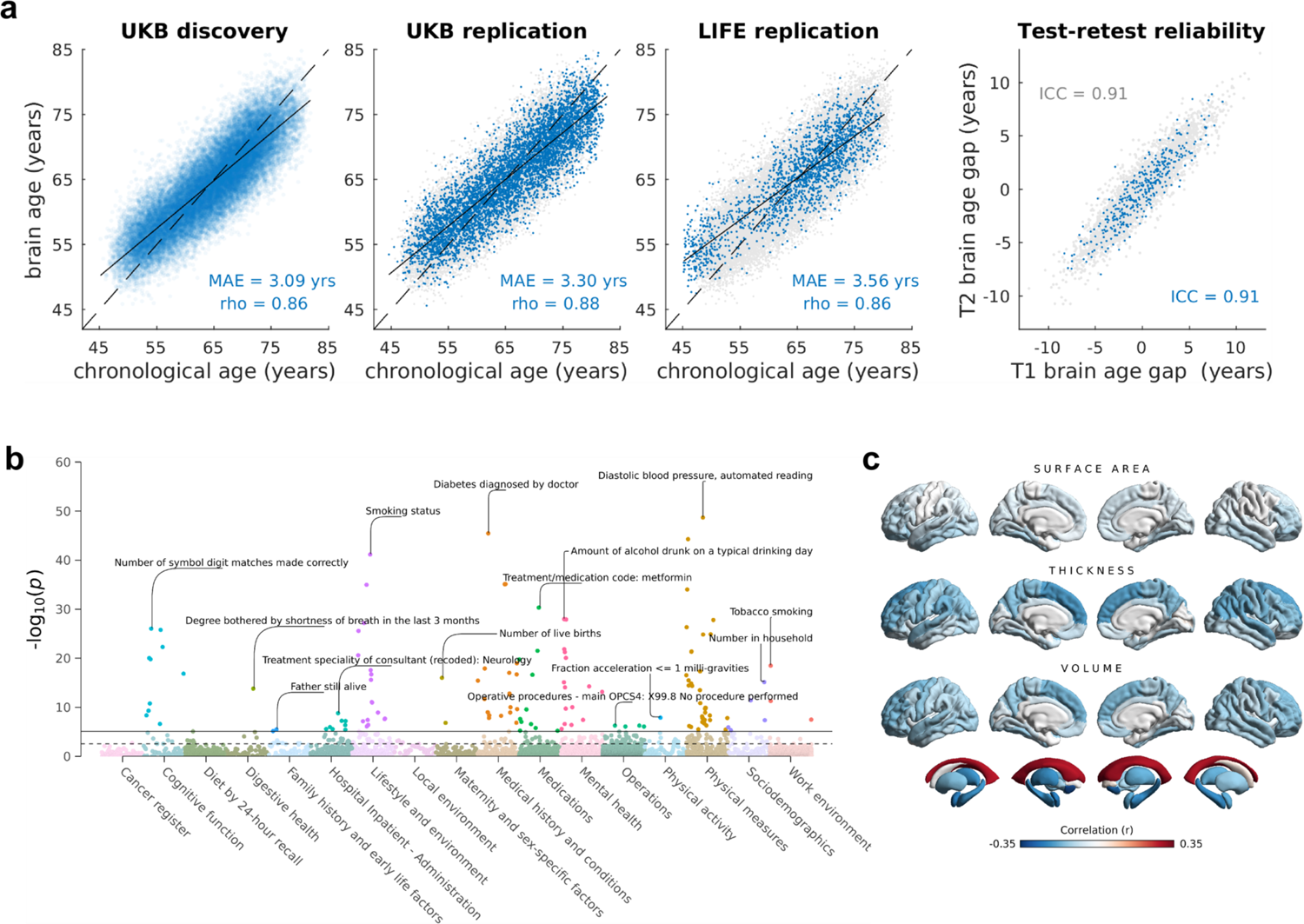
Prediction accuracies and phenotypic associations for combined grey and white matter BAG. **(a)** Blue dots in the first three plots (from left to right) show brain-predicted age estimates plotted against chronological age in the UKB discovery sample (*n* = 32,634), UKB replication sample (*n* = 5,427), and LIFE-Adult replication sample (*n* = 1,883). To facilitate comparisons, results of the UKB discovery sample are also shown as grey dots in the background of the UKB replication and LIFE replication plots. At this stage, brain-predicted age estimates have not yet been bias-corrected for regression dilution as indicated by the linear regression line (solid) crossing the identity line (dashed). The fourth plot shows the test-retest reliabilities of brain age gap in a subset of the UKB discovery (grey dots, *n* = 3,625) and UKB replication sample (blue dots, *n* = 376). Brain age gap was bias-corrected for age, age^2^, sex, scanner site, and total intracranial volume. **(b)** Cross-trait association results between brain age gap and 7,088 UK Biobank phenotypes from different health domains (sex, age, age2, scanner site, total intracranial volume served as covariates). Horizontal lines indicate the Bonferroni-adjusted (solid) and FDR-adjusted (dashed) level of significance. The top associations per category have been annotated. **(c)** Surface plots showing the correlations between brain age gap and 220 FreeSurfer brain structure variables. Colors reflect the strength and direction of partial product-moment correlations (sex, age, age^2^, scanner site, total intracranial volume served as covariates). MAE: mean absolute error; rho: product-moment correlation coefficient. ICC: intraclass correlation coefficient (C,1).^28^

**Table 1.** Prediction accuracies of the stacked age estimation models stratified by tissue class.

| | UKB discovery<br>($n = 32,634$; 45-82 years) | | | UKB replication<br>($n = 5,427$; 44-83 years) | | | LIFE replication<br>($n = 1,833$; 45-80 years) | |
| --- | --- | --- | --- | --- | --- | --- | --- | --- |
| | $r$ | MAE | ICC<sub>BAG</sub> | $r$ | MAE | ICC<sub>BAG</sub> | $r$ | MAE |
| Grey matter | .827 | 3.372 | .898 | .851 | 3.631 | .893 | .828 | 3.990 |
| White matter | .835 | 3.307 | .919 | .859 | 3.562 | .911 | .829 | 3.979 |
| Grey and white matter | .857 | 3.089 | .914 | .879 | 3.299 | .909 | .862 | 3.557 |

Note: Imaging data of the UKB discovery sample was released until January 2020 (release v1.7), while data of the UKB replication sample was released until September 2022 (release v1.9). $r$ : product-moment correlation between brain-predicted age (without bias correction) and chronological age; MAE: mean absolute error of brain-predicted vs. chronological age; ICC<sub>BAG</sub>: intraclass correlation coefficient between test and re-test assessment of brain age gap. Brain age gap was bias-corrected for age, age<sup>2</sup>, sex, scanner site, and total intracranial volume. ICCs are based on a subset of 3,625 subjects in the UKB discovery and 376 subjects in the UKB replication sample.

To validate our BAG estimates and examine phenotypic relationships with health-related traits, we conducted a cross-trait association analysis between BAG and 7,088 non-imaging-derived phenotypes using PHESANT.^29^ A total of 210 associations reached the Bonferroni-adjusted level of significance (*p* < 7.1e-06) for at least one of the three BAG traits (suppl. Fig. A2, suppl. Table B2). Fig. 1b shows the cross-trait association results for combined grey and white matter BAG.

The top associations for combined BAG (all *p* ≤ 1.8e-12) included ‘pack years of smoking’ (*r* = 0.091), ‘diastolic blood pressure, automated reading’ (*r* = 0.084), ‘number of symbol digit matches made correctly’ (*r* = −0.082), ‘diabetes diagnosed by doctor’ (*r* = 0.079), ‘amount of alcohol drunk on a typical drinking day’ (*r* = 0.076), and self-reported ‘overall health rating’ (*r* = 0.039; higher ratings indicate poorer health).

To explore how BAG is reflected in individual brain regions, we calculated BAG associations with FreeSurfer^30^ cortical surface measures and subcortical volumes (Fig. 1c, suppl. Fig. A3, suppl. Table B3). The strongest associations (all *p* ≤ 4.9e-146) were observed between BAG and volumes of the accumbentia (*r* = −0.31), lateral ventricles (*r* = 0.29), amygdalae (*r* = −0.25), and hippocampi (*r* = −0.22), as well as cortical thickness of the superior frontal (*r* = −0.16) and superior temporal cortex (*r* = −0.14). These results suggest that our models capture patterns of aging distributed throughout the brain, rather than being confined to specific brain areas. Moreover, results from cross-trait association analyses demonstrate relationships between BAG and multifaceted health traits, supporting the validity of BAG estimates.

### Discovery of 25 genomic loci

To identify genetic loci associated with BAG, we conducted GWAS analyses based on 9,669,404 Single Nucleotide Polymorphisms (SNPs) and insertion-deletions (INDELs) with MAF > 0.01 and INFO > 0.80. We modelled additive genetic effects, and used sex, age, age^2^, total intra-cranial volume, scanner site, type of genotyping array, and the first 20 genetic principal components as covariates. GWAS results for the three BAG traits are shown in Fig. 2.

**Fig. 2.**
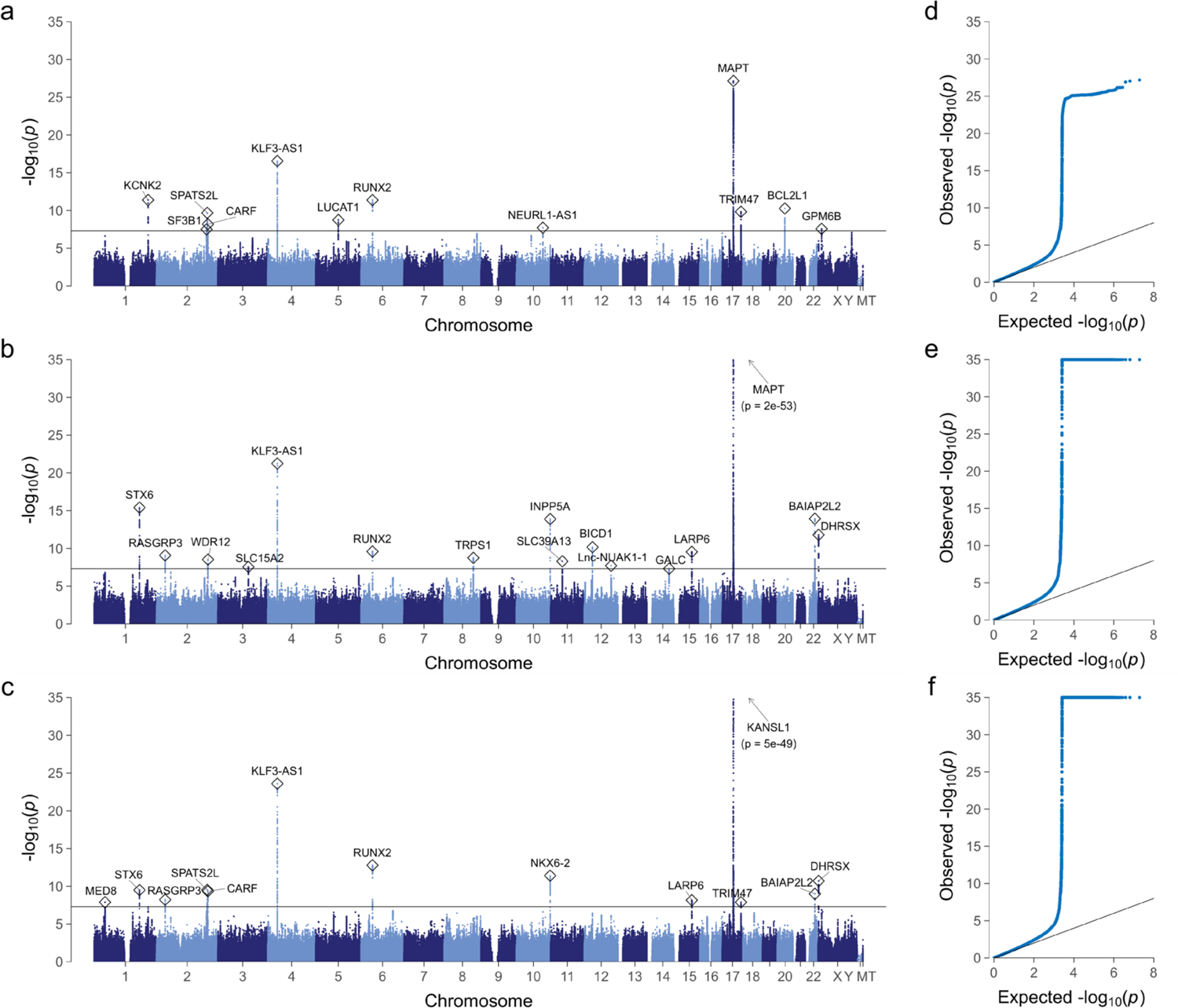
Manhattan plots (**a-c**) and quantile-quantile plots (**d-f)** showing the results of the discovery genome-wide association analyses for the three brain age gap traits (*N* = 32,634 UK Biobank individuals). Manhattan plots show the *p*-values (-log10 scale) of the tested genetic variations on the y-axis and base-pair positions along the chromosomes on the x-axis. The solid horizontal line indicates the threshold of genome-wide significance (*p* = 5E-8). Index variations are highlighted by circles and were annotated with those genes implicated by our gene prioritization analysis. Results of pseudoautosomal variations have been added to chromosome ‘X’. Quantile-quantile plots show the observed *p*-values from the association analysis vs. the expected *p*-values under the null hypothesis of no effect (-log10 scale). For illustrative reasons, the y-axis has been truncated at *p* = 1e-35. **a,d** grey matter brain age gap; **b,e** white matter brain age gap; **c,f** combined grey and white matter brain age gap.

LD score regression (LDSC) intercepts did not indicate a bias of test statistics due to reasons other than polygenicity, suggesting no confounding inflation due to population stratification (intercept range: 1.0075-1.0142; suppl. Table B4).^31^ SNP-based heritability estimates derived from LDSC ranged between 26.2% (grey matter BAG) and 28.6% (white matter BAG). Estimates from GCTA-GREML^32^ were slightly higher, suggesting SNP-based heritabilities of 28.9% for grey matter, 32.7% for white matter, and 32.3% for combined grey and white matter BAG (*SE*: 1.3%). The bivariate genetic correlation between grey and white matter BAG reached *r_G_* = 0.703 (*SE* = 0.018), suggesting both shared and distinct genetic contributions. Stratified LDSC (suppl. Fig. A4, suppl. Table B5) revealed an enrichment of heritability (all *FDR* < 0.05) in regions evolutionary conserved across mammals (fold enrichment *FE*: 13.9) and primates (*FE*: 12.7). We also observed heritability enrichment in super enhancer (*FE*: 2.7), and epigenetically modified H3K27ac (*FE*: 2.0), H3K4me1 (*FE*: 1.9), and H3K9ac regions (*FE*: 2.9).

To identify independent genome-wide significant associations, we conducted stepwise conditional analyses using GCTA-COJO.^32,33^ This resulted in 12 independent discoveries for grey matter BAG, 16 for white matter BAG, and 13 for combined BAG (regional plots shown in suppl. Fig. A5-12). Across the three BAG traits, the total count of independent discoveries was 25 (Table 1; suppl. Table B6), as determined through cross-trait LD clumping of index variations (R^2^ > 0.1; 10 Mbp window-size). These 25 loci represent distinct genomic regions with a physical distance larger than 2.5 Mbp. Among the 25 loci, 12 have previously been reported genome-wide significant for BAG,^15,17–20^ thus, 13 loci are novel findings.

We observed the majority of index variations in intronic regions of protein-coding genes. Consistently, ANNOVAR enrichment tests indicated that variants in high linkage disequilibrium (LD) with our genome-wide significant variants were underrepresented in intergenic regions, and over-represented in UTR3, UTR5, exonic, intronic, exonic non-coding RNA, and intronic non-coding RNA regions (suppl. Fig. A13, suppl. Table B7).

### Fine-mapping and gene prioritization

To shed light on the potential causal genes through which identified variants exert their effects on BAG, we used several fine-mapping, annotation, and co-localization strategies that integrate information from multiple omics resources. For each genome-wide significant locus, we a) inferred the number of distinct causal signals and constructed 95% credible sets of variants that likely include the causal variant using FINEMAP,^34^ b) physically mapped credible variants to genes using ANNOVAR,^35^ c) predicted transcript consequences of non-synonymous exonic credible variants and scored their deleteriousness using CADD,^36^ DANN,^37^ and REVEL,^38^ d) mapped variants to genes through expression quantitative trait locus (eQTL) lookup in 49 GTEx v8 tissues,^39^ e) conducted summary-data-based Mendelian Randomization (SMR)^40,41^ with RNA sequence data of 2,865 brain cortex samples^42^ to test for mediation of variant effects through gene expression and splicing, and f) calculated polygenic priority scores (PoPS)^43^ that incorporate data from single-cell RNA sequencing datasets, curated biological pathways, and protein-protein interaction (PPI) networks. Across all genes nominated by abovementioned strategies, we computed an integrative gene priority sore and prioritized the most relevant gene (see methods). Figure 3 provides an overview of the analysis workflow. A locus-wise summary of all results is shown in suppl. Table B6 (details in suppl. Tables B7-B15).

**Fig. 3.**
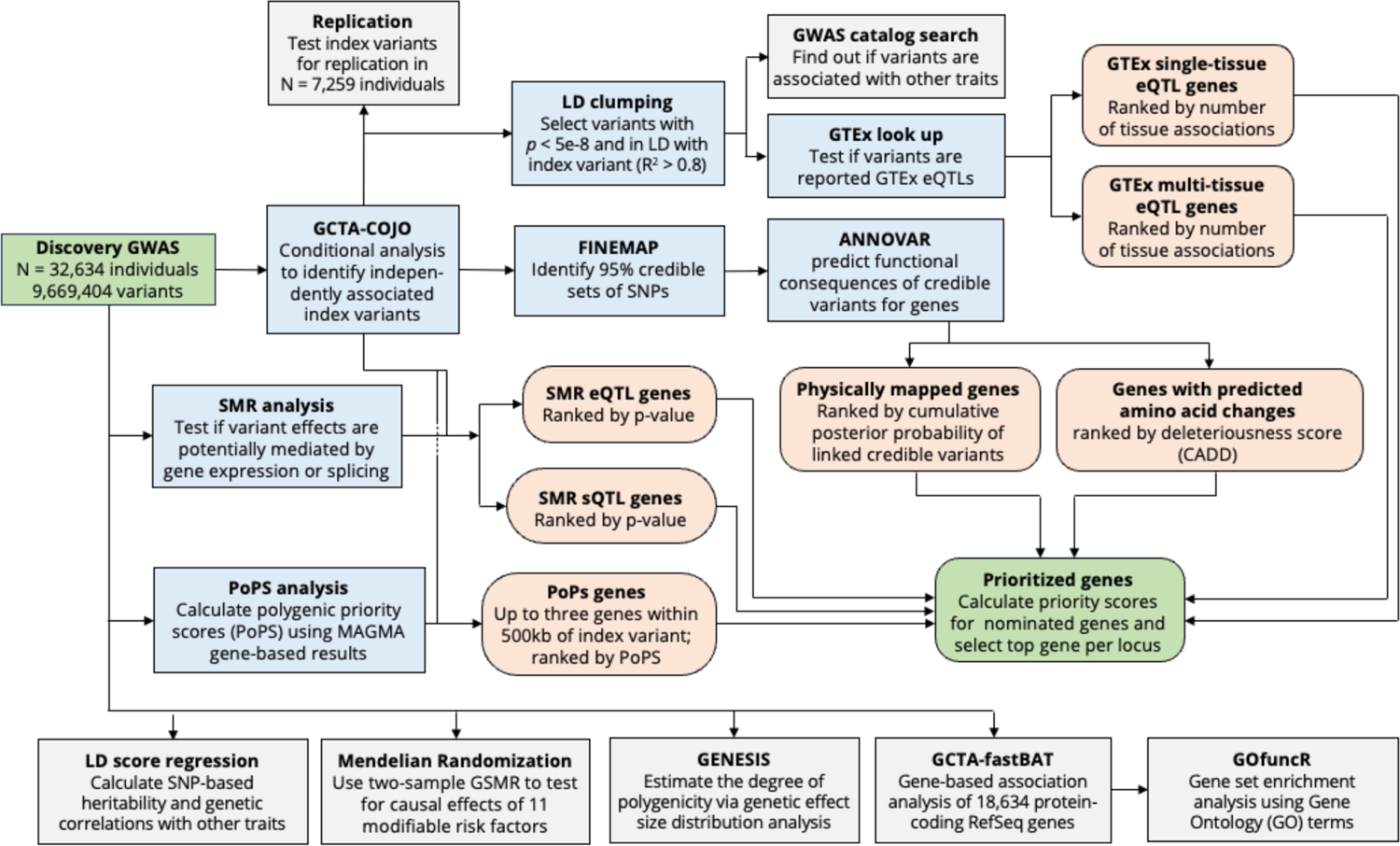
Overview of the post-GWAS analysis workflow including the gene prioritization procedure. Green boxes represent data input (discovery GWAS) and output (prioritized genes). Blue boxes represent analyses whose outcomes were used for gene nomination and subsequent prioritization. Apricot-colored boxes reflect gene nomination categories. Grey boxes reflect all other analyses carried out to refine the genetic architecture of brain age gap such as heritability and polygenicity analyses. Genes were prioritized by integrating data from multiple strategies such as functional annotation of credible variants, summary-data-based Mendelian Randomization (SMR), GTEx eQTL lookups, and Polygenic Priority Scores (PoPS).

For the 25 discovered loci, FINEMAP revealed a model-averaged number of *k* causal signals per locus ranging from 1.04 to 2.06 (median: 1.24), with the most probable *k* model suggesting 1 causal signal for 21 loci, and 2 causal signals for 4 loci (suppl. Table B6). This finding is largely consistent with conditional analysis results, suggesting 1 independent signal for each locus. The size of the 95% credible set of variants ranged between 4 and 2,514 (median: 46), indicating a small pool of causal candidates for some loci and a putative complex linkage structure that hinders pinpointing causal variants for other loci. The estimated per-locus contribution to the phenotypic variance, i.e., the regional heritability, ranged between 0.09% and 0.70%.

We observed the strongest associations at locus 17q21.31 (rs111854640, *p* = 2.3E-53), which tags a well-known 900kb inversion polymorphism.^44,45^ This region is one out of three with alternate haplotype reference sequences included in genome assembly GRCh37 (UCSC haplotype sequence: chr17_ctg5_hap1). Consistent with the strong LD cluster in the inverted region,^44^ we found this locus with by far the largest credible set of variants (2,514). We carried out an NHGRI-GWAS catalog search to identify pleiotropic effects with other complex traits.^46^ This search revealed a large variety of locus-associated traits, including educational attainment,^47^ depressed affect,^48^ alcohol consumption,^49^ sleep duration,^50^ lung function,^51^ male puberty timing,^52^ age at onset of menarche,^51^ and Alzheimer’s disease.^53^ The locus covers multiple genes, including *MAPT, STH*, *KANSL1*, and *CRHR1*. Several genome-wide significant variants in these genes are GTEx single-tissue and multi-tissue eQTLs (suppl. Tables B12-B13). SMR analyses implicated alterations in gene expression and splicing of *MAPT* and *KANSL1* as putative mechanisms that mediate variant effects on BAG (suppl. Tables B10-11). Additionally, we identified several exonic variants causing amino-acid changes in transcript sequences (suppl. Table B9). The highest CADD deleteriousness score was shown for rs176515149 (CADD score: 34), located in exon 6 of *MAPT*, with a GWAS *p*-value of 4.9e-52. This variant causes an arginine-to-tryptophan substitution at MAPT protein position 370. *MAPT* encodes the well-known ‘tau protein’ implicated in Alzheimer’s and other neurodegenerative diseases.^54^ Altogether, we prioritized *MAPT* as most plausible susceptibility gene for brain aging in this locus.

Three other loci were identified with a tractable number (≤ 10) of likely causal variants. The first locus with 4 credible variants refers to an intergenic region at 1q41, 41 kb upstream of *KCNK2*, encoding Potassium channel subfamily K member 2. *KCNK2* is also prioritized by SMR, GTEx, and PoPS analyses. *KCNK2* has been implicated in neuroinflammation, blood-brain barrier dysfunction, and cerebral ischemia.^55,56^. GWAS catalog matches suggest associations with cortical thickness,^57^ surface area,^58^ and sulcal opening.^59^.

The second locus, again with 4 variants in the 95% credible set, refers to an intronic region of NUAK1 at 12q23.3. Index variant rs12146713 is a multi-tissue eQTL of Lnc-NUAK1-1, i.e., a long non-coding RNA gene expressed in brain cortex, cerebellum, and other tissues. GWAS catalog matches suggest further associations with cortical thickness,^57^ surface area,^58^ and subcortical volume.^57^

The third locus refers to exon 11 of *BAIAP2L2* at 22q13.1, with 9 variants in the 95% credible set. Index variant rs142739979 is a non-frameshift INDEL predicted to cause an insertion of threonine, proline, and methionine between BAIAP2L2 protein sequence positions 411 and 412. Additionally, this variant is a reported eQTL of nine genes, including *SLC16A8* linked to age-related macular degeneration,^60^ and *TRIOBP*, whose protein isoforms have been implicated in neurite outgrowth, cell cycle progression, and motility of cancer cells.^61^

While above-mentioned loci have also been supported in previous studies, we here discover several novel loci that offer new insights into the biological path-mechanisms of brain aging. One of them refers to a UTR5-region of *TRPS1* at 8q23.3, with *TRPS1* also representing the prioritized gene. Previous studies have implicated variants in strong LD (R^2^ > 0.8) with the index variant in neuroticism,^48^ type-2 diabetes,^62^ and anthropometric measures.^63^ TRPS1 is a transcription factor that represses the expression of GATA-regulated genes in vertebrate development,^64^ and has been implicated in a large variety of physiological processes including organ differentiation and tumorigenesis.^65^

Another novel discovery refers to an intronic region of *ICA1L* at 2q33.2, led by rs76122535 (*p* = 4.6e-10), with GTEx and SMR analysis indicating variant effects on expression and splicing of *ICA1L*, *CARF*, and *NBEAL1* in a variety of tissues. The highest priority score was attained by *CARF*, which is reportedly upregulated during stress-induced and oncogenic senescence, and its overexpression has been observed to cause premature senescence.^66^

Furthermore, we identified an X-chromosomal locus in an intron of *GPM6B,* led by rs597999 (*p* = 2.7e-08), which is also a GTEx eQTL of *GPM6B*. The GPM6B protein has been suggested to regulate serotonin uptake, is particularly expressed in brain tissue, and belongs to the proteolipid protein family involved in cell-to-cell communication.^67^

A final novel discovery to be mentioned refers to a variant near *DHRSX*, tagged by rs377113838 (*p* = 1.6e-12), which is a GTEx multi-tissue eQTL of the same gene, and lies in a pseudoautosomal region, i.e., a region with homologous sequences on chromosome X and Y. DHRSX has been shown to be play a crucial role in starvation-induced autophagy.^68^

As strongest known risk factor for Alzheimer’s disease, we tested the ε4 allele of the apolipoprotein E (*APOE*) gene, determined from haplotypes of rs429358 and rs7412, for BAG associations. The number of ε4 alleles was indeed associated with higher BAG (*p* = 7.8e-06), although not attaining genome-wide significance.

Altogether, by integrating information from several fine-mapping, functional annotation, and colocalization strategies we here prioritized several genes potentially involved in brain aging. These leads may stimulate novel testable hypotheses on the causes of biological aging.

### Replication of discovered variants

Independent discoveries from the SNP-level analysis were tested for replication in a follow-up cross-ancestry meta-analysis of up to 7,259 individuals (suppl. Table B15, suppl. Fig. A14-A17). Replication analyses included index variations from the 25 genome-wide significant loci, and index variations from another 45 suggestive loci (conditional *p*-values ranging from 1.0e-06 to 5.0e-08). The degree of consistency between discovery and replication results was highly unlikely to occur by chance (suppl. Fig. A18). Of the 25 discoveries, 23 showed consistent effect directions (binomial test: *p* = 9.7e-06) and 16 replicated at *p* < 0.05 (one-tailed nominal significance; binomial test: *p* = 2.0-15). This finding aligns closely with the outcomes predicted by statistical power analyses, with 16.51 out of 25 loci expected to attain one-tailed nominal significance in replication analyses. All replicated loci are highlighted in Table 2 (column ‘Rep.’). Novel replicated loci included 2q33.2 (rs76122535 near *CARF*), 3q13.33 (rs34567530 in *SLC15A2*), 5q14.3 (5:90567689_TTA_T near *LUCAT1*), 8q23.3 (rs2721939 in TRPS1), 17q25.1 (rs3833085 in *TRIM47*), Xp22.2 (rs5979992 in *GPM6B*), and Xp22.33 (rs377113838 near *DHRSX*, pseudoautosomal).

**Table 2.**
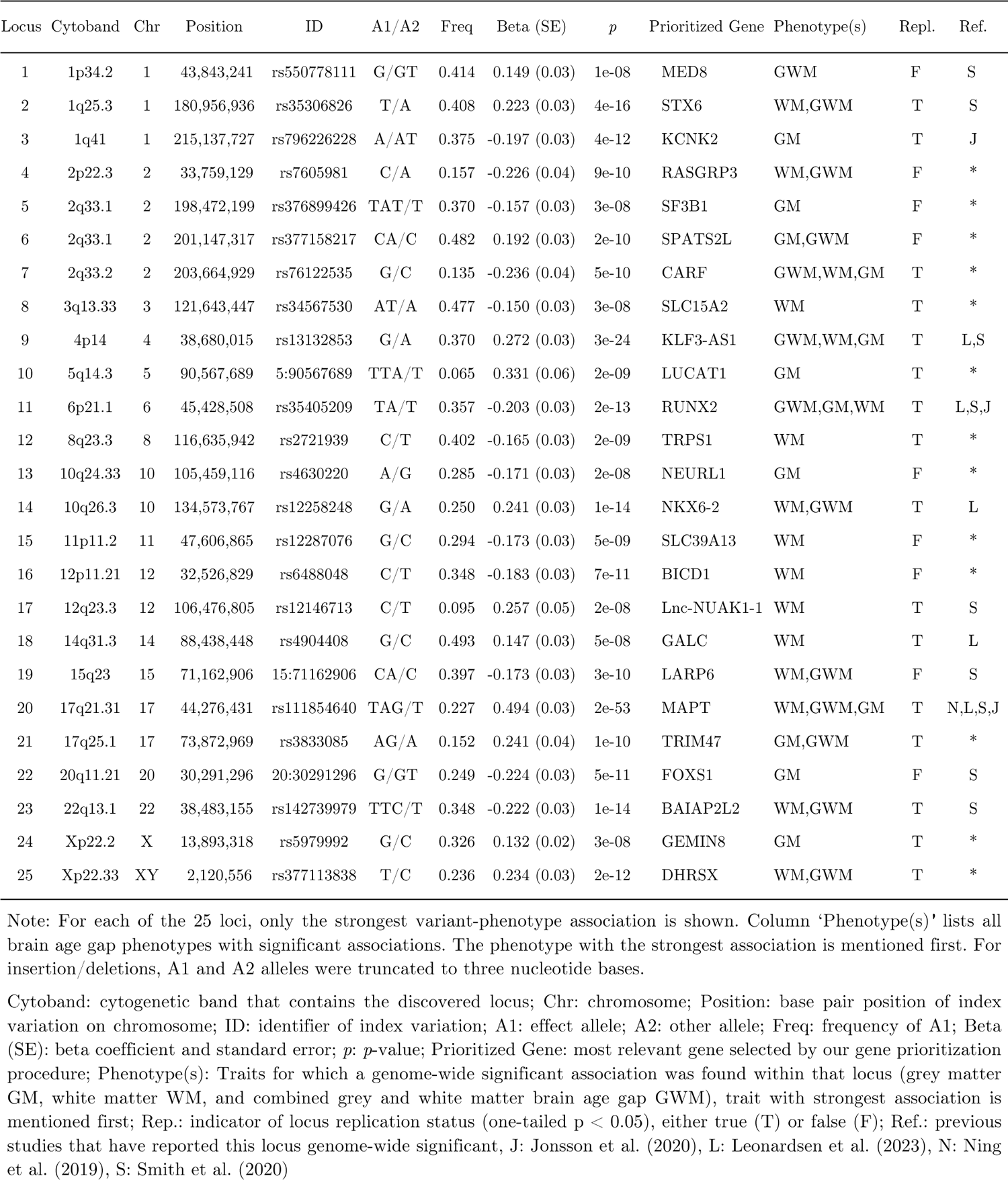
Independent loci discovered through GWAS analyses of brain age gap in *N* = 32,634 individuals.

Among the additional 45 suggestive variants, we found 35 with consistent effect directions (binomial test: *p* = 1.2e-04) and 14 attaining one-tailed nominal significance in replication analyses (binomial test: *p* = 2.3e-08). Moreover, we found polygenic scores (PGS) with variance explanations of 2.1% for grey matter, 2.8% for white matter, and 2.5% for combined BAG (all *p* ≤ 5.1e-22; suppl. Table B17). Proportions of explained variance were lower when considering only genome-wide significant discoveries (0.8-1.3%; all *p* ≤ 1.5e-09). In sum, replication results provide strong support for true associations among the discovered loci and point towards additional contributions at sub-threshold significance levels.

### Gene-based analysis

We investigated the potential impact of 18,643 protein-coding genes using GCTA-fastBAT.^69^ Gene-based analyses aggregate information from multiple variants within the same genomic region, resulting in a reduced multiple-testing burden compared to variant-based analyses. Gene-based analyses revealed 188, 327, and 295 genes significantly associated (FDR < 0.05) with grey matter, white matter, and combined grey and white matter BAG, respectively. To identify independent associated loci, we conducted a *p*-value informed clumping procedure of genes located in a physical distance of 3,000 kbp. This resulted in 69, 114, and 97 distinct loci, respectively, of which 151 were unique across the three phenotypes (suppl. Fig. A19, suppl. Table B18). Again, the strongest signal was observed at 17q21.31 covering *MAPT*. Significant genes also included *APOE* (encoding apolipoprotein E), i.e., the gene with the strongest known impact on late-onset Alzheimer’s disease.^70^ In total, gene-based analyses provide evidence for an extended set of genomic loci involved in human brain aging.

### Pathway analysis

To gain further insights into the biological mechanisms underlying brain aging, we used gene-based results to test for an enrichment of Gene Ontology (GO) terms, i.e., sets of genes known to serve a common biological function.^71^ Gene set enrichment analyses (GSEA) were conducted using GOfuncR.^72^ GSEA revealed nine significant GO terms (suppl. Table B19) after refinement of hierarchical dependencies. Analyses provided indications for a role of the immune system in brain aging, with significant results obtained for ‘MHC protein complex’ (GO:0042611) and ‘peptide antigen binding’ (GO:0042605). Results also implicated ‘small GTPase binding’ (GO:0031267l) as potential mechanism in brain aging. Small GTPases are a superfamily of evolutionary conserved proteins that act as biological timers (binary on/off switches) of many essential cellular processes.^73,74^ These processes include cell differentiation, proliferation, and signal transduction.^75^ Several small GTPase proteins have been implicated in premature senescence.^76–78^

### Genetic correlations with other complex traits

To examine a potential shared genetic basis with other complex traits, we applied bivariate LD score regression^31,79^ to GWAS summary statistics and calculated genetic correlations with 38 frequently employed traits from different mental and physical health domains (suppl. Table B20).^80–82^ We also calculated genetic correlations with 989 heritable traits from a large set of GWAS summary statistics (Zenodo: https://doi.org/10.5281/zenodo.7186871). In total, we observed 22 out of 38 selected traits to significantly correlate (FDR < 0.05) with at least one of the three BAG variables (Fig. 4a, suppl. Table B21). Grey matter BAG showed the highest number of significant associations (22), relative to white matter (1) and combined grey and white matter BAG (13). A similar pattern was observed for the 989 traits, where we observed 121, 2, and 36 significant associations for the three BAG traits, respectively (Fig. 4b and 4c, suppl. Table B22).

**Fig. 4.**
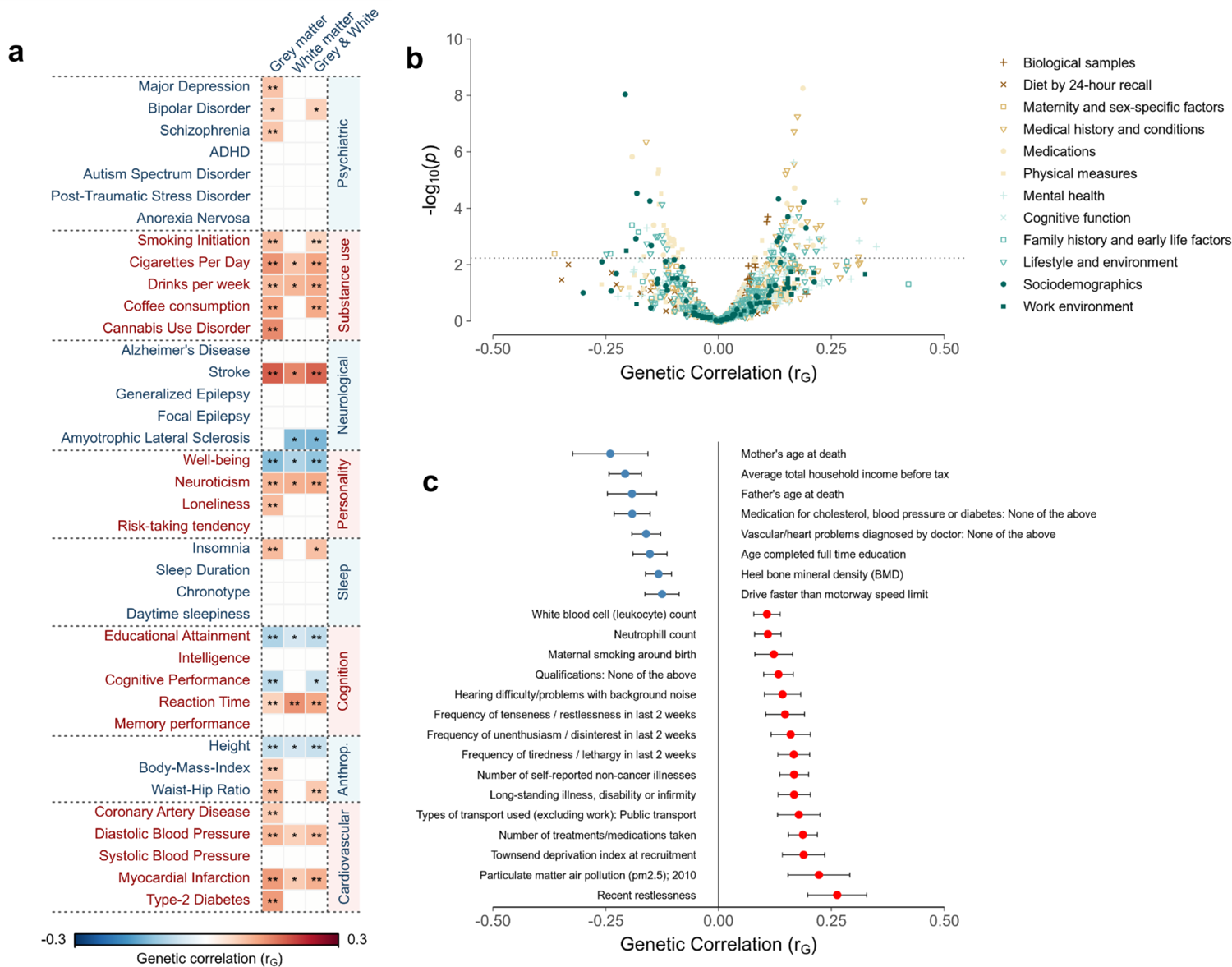
Results from the genetic correlation analyses between brain age gap and 1,027 other complex phenotypes. **(a)** Genetic correlation matrix between brain age gap (columns) and 38 selected phenotypes from different health domains (rows). * *p* < 0.05 (nominal significance) ** FDR < 0.05 (level of significance after multiple testing-correction) (**b)** Volcano plot showing the magnitude (x-axis) and significance (y-axis) of genetic correlations between grey matter brain age gap and 989 traits examined by Neale and colleagues. The dashed horizontal line indicates the FDR-adjusted level of significance. (**c)** Forest plot showing the genetic correlation coefficients and standard errors for a subset of 23 exemplary traits that showed significant genetic correlations with grey matter brain age gap.

Among the 38 selected traits, we found significant associations between grey matter BAG and psychiatric (e.g., Major Depression: *r*_G_ = 0.085), substance use (cigarettes per day: *r*_G_ = 0.134), neurological (stroke: *r*_G_ = 0.182), personality (neuroticism: *r*_G_ = 0.100), sleep-related (insomnia: *r*_G_ = 0.096), cognition-related (educational attainment: *r*_G_ = −.091), anthropometric (body-mass-index: *r*_G_ = 0.075), as well as cardiovascular and metabolic syndrome traits (type-2 diabetes: *r*_G_ = 0.127).

Among the 989 heritable traits, we found evidence for genetic correlations with ‘mother’s age at death’ (*r*_G_ = −0.240) and ‘father’s age at death’ (*r*_G_ = −0.192), suggesting that higher BAG is associated with shorter familial life expectancy. Other significant associations referred to socioeconomic variables (e.g., ‘average total household income before tax’: *r*_G_ = −0.207), mental health variables (‘frequency of tiredness/lethargy in last 2 weeks’: *r*_G_ = 0.167), medical conditions (‘Vascular/heart problems diagnosed by doctor: High blood pressure’: *r*_G_ = 0.149), medication intake (e.g. ‘medication for cholesterol, blood pressure or diabetes: None’: *r*_G_ = −0.191), early life factors (‘maternal smoking around birth’: *r*_G_ = 0.122), among others (suppl. Table B22). These results suggest a shared genetic basis between BAG and a broad range of health-related variables.

### Causal associations

We used two-sample generalized summary-data-based Mendelian Randomization (GSMR)^83^ to investigate potential causal effects of 12 modifiable risk/resilience factors on BAG. The risk/reliance factors were BMI, waist-hip-ratio adjusted for BMI, low-density lipoprotein cholesterol (LDL-c), high-density lipoprotein cholesterol (HDL-c), triglycerides, systolic blood pressure, diastolic blood pressure, pulse pressure, type-2-diabetes, coronary artery disease, schizophrenia, and years of education. Our analyses revealed significant effects of diastolic blood pressure on all three BAG traits (combined BAG: *β*_xz_ = 0.610, *p* = 1.4e–08), as well as systolic blood pressure on grey matter (*β*_xz_ = 0.443, *p* = 5.8e-05) and combined BAG (*β*_xz_ = 0.326, *p* = 0.002).^84^ Results suggest that one standard deviation increase in blood pressure causally contributes to an about half-year increase in BAG (suppl. Table B24, suppl. Fig. A20-22).

### Degree of polygenicity and projection of discoveries to future GWAS

To quantify the degree of polygenicity and predict the number of discoveries in forthcoming GWAS, we used GENESIS^85^ and estimated the number of underlying susceptibility variants and their effect sizes. We considered ‘height’ and ‘neuroticism’ as benchmark traits due to their different degrees of polygenicity.^86–89^ For the three BAG traits, the number of susceptibility variants was consistently estimated at 5.7k (SE 1.7k; suppl. Table B25). By comparison, this number was estimated at 12.6k (SE 1.3k) for height and 16.2k (SE 1.2k) for neuroticism. The distributions of variant effect sizes (Fig. 5a) revealed that BAG exhibits a greater proportion of contributing variants with large effect sizes when compared to neuroticism and standing height.

**Fig. 5.**
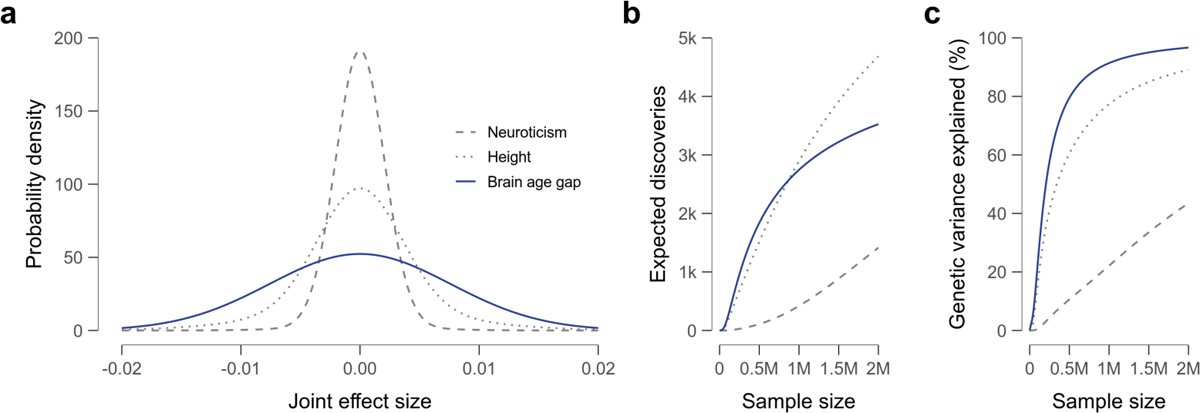
Results of the genetic effect size distribution analysis for combined grey and white matter brain age gap. Neuroticism and standing height serve as reference traits. (**a)** Effect-size distributions of underlying susceptibility variants. Wider tails indicate a greater proportion of susceptibility variants with large effect sizes. (**b)** Expected number of discoveries as a function of sample size. (**c)** Expected proportion of genetic variance explained by genome-wide significant discoveries as a function of sample size.

Moreover, the number of discoveries for BAG was predicted to show rapid increases with rising sample sizes (Fig. 5b). Fig. 5c shows that about 500k subjects are required to explain 80% of the SNP-based heritability for BAG from genome-wide significant discoveries. This number aggregates to about 1M for height and 6M for neuroticism. Together, these results suggest a relatively low degree of polygenicity for BAG when compared to other complex traits.

## Discussion

We here leveraged genomic and neuroimaging methods to demonstrate the significance of brain age gap (BAG) as a putative biomarker of aging and its prospective utility in identifying therapeutic targets. Machine learning and MRI quantified brain age with excellent measurement properties, capturing distributed patterns of aging across the brain. Cross-trait association analyses established robust associations with various health traits, highlighting the potential clinical relevance of BAG. We showed that BAG is under substantial genetic control, with about 30% of the phenotypic variance attributable to common genetic variation. We identified 25 independent genome-wide significant loci, of which 13 loci are novel. The observed genomic signals unveiled several enriched biological pathways, e.g., immune-system-related processes as well as the binding of small GTPases, prompting further mechanistic exploration. Using genetic correlations, we characterized the common genetic basis between BAG and other complex traits, including psychiatric, neurological, cognitive, personality, substance use, sleep-related, as well as cardiovascular and metabolic syndrome traits. Through Mendelian Randomization, we established evidence for a causative role of enhanced blood pressure in accelerated brain aging.

Finally, we find BAG with a relatively low degree of polygenicity, and we anticipate this will facilitate further variant discoveries in the near future. Brain aging is not a uniform process; rather, it encompasses diverse aspects of structural and functional change. Studying different aspects of brain aging has been advocated to increase the yield of biologically meaningful insights.^18^ We calculated separate BAG for the brain tissue classes grey matter and white matter, in addition to a composite measure. For both tissues, we found remarkably consistent age prediction accuracies, along with comparable test-retest reliabilities and heritabilities. The genetic correlation between grey and white matter BAG, however, settled at r_G_ = 0.70 (SE 0.018), suggesting both shared and segregated biological underpinnings. While grey matter and white matter BAG exhibit comparable SNP-based heritabilities (0.289 vs. 0.327; SE: 0.013), grey matter BAG showed a noticeably higher number of significant phenotypic and genetic associations, which may imply greater relevance for several health dimensions. This subject warrants more in-depth exploration in future research.

We confirmed the previously reported inversion locus at 17q21.31 as strongest known genetic contributor to BAG,^17,19–21^ explaining 0.3% of the variance in grey matter and 0.7% in white matter BAG. The most likely causative gene in this locus, *MAPT*, encodes the well-known ‘tau’ protein associated with Alzheimer’s disease. Genomic analyses also unveiled a role of Alzheimer’s risk gene *APOE* and other apolipoprotein genes. With both tau- and apolipoprotein alterations, our results implicate two hallmarks of Alzheimer’s in accelerated brain aging. This aligns with the demonstrated capability of BGA to forecast Alzheimer’s disease.^90^

Two prior studies on BAG have attempted to replicate variant discoveries, albeit with limited success.^17,20^ Here we observed a high degree of consistency between discovery and replication results, with 23 out of 25 loci showing consistent effect directions, and 16 loci attaining nominal significance in replication analyses. Notably, polygenic scores accounted for about 2-3% of the phenotypic variance, a remarkable proportion when compared to traits such as intelligence and major depressive disorder, which necessitated considerably larger discovery samples to attain similar prediction accuracies.^82,91^

The current study has several limitations. First, the employed gene prioritization techniques face challenges in pinpointing causal genes,^43^ particularly for loci characterized by high gene density and complex linkage structures. Second, BAG has been estimated from cross-sectional data, which is typically interpreted as accelerated or decelerated aging. However, an alternative interpretation posits BAG as stable individual differences that emerge at an ontogenetically early period and are carried into old age.^92^ Third, statistical power analyses revealed an expected number of 16 successful replications (out of 25), indicating a need for higher replication sample sizes. Fourth, polygenic overlap between different traits was estimated using genetic correlations. Yet, this technique does not capture fractions of genetic variants shared between two traits irrespective of variant effect directions. Other bioinformatic tools such as MiXeR may be used in future studies to quantify genetic overlap by considering mixtures of variant effects.^93^ Fourth, polygenicity models are known to classify variants with very low effect sizes as null, resulting in a likely underestimation of BAG polygenicity estimates.

In conclusion, the present study refines the genetic architecture of brain age gap and its relationships to other traits. We added 13 new variants to the catalogue of existing BAG associations and assigned plausible candidates to these loci such as TRPS1 implicated in various pathological processes, GPM6b involved in cell-to-cell communication, and CARF linked to premature senescence. This will facilitate further work on path-mechanisms of BAG and potential therapy targets.

## Data Availability

The individual-level data incorporated in this work have been obtained from the UK Biobank resource (https://www.ukbiobank.ac.uk/). GWAS summary statistics for BAG will be made available at Zenodo (https://zenodo.org/) upon publication of this article.

## Acknowledgments

This research has been conducted using the UK Biobank Resource under Application Number 423032. This publication was supported by LIFE – Leipzig Research Centre for Civilization Diseases, University of Leipzig. LIFE was funded by means of the European Social Fund and the Free State of Saxony. We thank the ICBP consortium for providing summary statistics.

## Code availability

All scripts used in this work are available on GitHub (https://github.com/pjawinski/ukb_brainage)

## Supplementary Material

Supplementary information is available at bioRxiv online.

## Methods

### Sample characteristics

Participants were drawn from the UK Biobank cohort study (www.ukbiobank.ac.uk) under application number 423032. A detailed description of the UK Biobank study design, participants and quality control (QC) methods has been published previously.^23^ The UKB received ethical approval from the National Research Ethics Service Committee North West-Haydock (reference 11/NW/0382). All participants provided written informed consent. In the current study, the majority of participants were drawn from the January 2020 UKB brain imaging release (v1.7). These data contained 40,681 participants with structural T1-weighted MRI data (UKB data-field 20252). We did not include T1-weighted MRI scans in folders labelled as ‘unusable’ (leaving 39,679 participants). In total, MRI scans of 39,677 participants completed the voxel-based morphometry preprocessing (see section ‘MRI acquisition and preprocessing’). Analyses were restricted to participants whose self-reported sex matched the genetic sex (data-field 31 and 2200), who were without indications of sex aneuploidy (data-field 22019), and who were no outliers in heterozygosity and missingness (data-field 22027). We only included unrelated participants as suggested by pairwise kinship coefficients below 0.0442 (pre-calculated coefficients retrieved using the command line tool ‘ukbgene’ with the ‘rel’ parameter). In our discovery GWAS, we only included participants of white-British ancestry (data-field 22006), which resulted in a final discovery sample of 32,634 participants (17,084 female, age range: 45.2-81.9 years, mean age: 64.3 years).

For replication analyses, we selected all remaining non-white-British ancestry individuals of the January 2020 release. Applying the same inclusion criteria, we then added participants whose imaging data were released until September 2022 (release v1.9), yielding a total of 7,785 additional individuals. None of these individuals were related to individuals in the discovery sample. We only included individuals with a valid ancestry assignment from the Pan-ancestry UKB project (UKB return 2442; https://pan.ukbb.broadinstitute.org/). This resulted in 217 African, 60 Admixed American, 409 Central/South Asian, 192 East Asian, 4,486 European, and 62 Middle Eastern ancestry participants. In total, we included 5,427 UKB participants for replication analyses (2,847 female, age range: 44.6-82.8 years, mean age: 65.9 years). From the LIFE-Adult cohort study,^26,27^ we included another 1,833 European ancestry participants (888 female, age range: 45.2-80.3 years, mean age: 65.3 years).^26,27^ Altogether, the final replication sample included 7,259 participants (3,735 female, age range: 44.6-82.8 years, mean age: 65.7 years) from 7 subsamples (see section ‘Genome-wide association analysis and replication meta-analysis meta-analysis’ for analysis details).

### MRI data acquisition

The UKB imaging acquisition protocol and processing pipeline have been detailed previously (http://biobank.ctsu.ox.ac.uk/crystal/refer.cgi?id=1977). In brief, brain MRI data were acquired in one of four UKB imaging centers (Cheadle, Newcastle, Reading, and Bristol) on Siemens Skyra 3T MRI scanners (Siemens Healthcare, Erlangen, Germany), running software VD13A SP4 with a standard Siemens 32-channel RF receive head coil. UKB’s neuroimaging strategy includes the acquisition of several imaging modalities. In this study, we used T1-weighted structural MRI scans acquired using a 3D MPRAGE sequence in the sagittal plane, with 1×1×1 mm voxel-size, 208×256×256 acquisition matrix, 2,000 ms repetition time (TR), 2.01 ms echo time (TE), 880 ms inversion time (TI), 6.1 ms echo spacing, 8° flip angle, 240 Hz/pixel bandwidth, in-plane acceleration factor of *R* = 2, and 4:54 min duration (https://www.fmrib.ox.ac.uk/ukbiobank/protocol/). T1-weighted scans were defaced for subject anonymity and made available in NIFTI format (data-field 20252).

In LIFE-Adult, brain imaging was performed using a 3T Siemens Verio MRI scanner (Siemens Healthcare, Erlangen, Germany) equipped with a standard 32 channel head coil. High resolution T1-weighted structural images were obtained using a 3D MPRAGE sequence with 1×1×1 mm voxel-size, 256×240×176 acquisition matrix, TR = 2,300 ms, TE = 2.98 ms, TI = 900 ms, and 9° flip angle.

### MRI preprocessing

T1-weighted MRI scans in NIFTI-format were preprocessed using the voxel-based morphometry pipeline of CAT12 (r1364, http://dbm.neuro.uni-jena.de) for SPM12 (r7487) in MATLAB R2021a (The MathWorks Inc, Natick, MA, USA). CAT12 preprocessing involved affine and DARTEL registration of brain images to a reference brain, segmentation into grey matter, white matter, and cerebro-spinal fluid, bias correction for intensity inhomogeneity, and modulation of segmentations to account for the amount of volume changes due to spatial registration. Processed images were smoothed by applying an 8×8×8mm full-width-at-half-maximum (FWHM) gaussian kernel with subsequent resampling to 8mm^3^ voxel size. We only considered MRI scans with a CAT12 overall image quality rating < 3.0 for further downstream analyses.

### Feature set for machine learning

The feature set for machine learning was derived from CAT12-preprocessed grey and white matter segmentations, with the smoothed and resampled brain images comprising 16128 voxels each. We excluded voxels that did not show any variation across individuals, resulting in 5416 voxels for grey matter images and 5123 voxels for white matter images. Typically, voxel-based images are characterized by substantial spatial correlations. We conducted principal component analysis (PCA) in MATLAB to remove redundant information and reduce dimensionality. We selected the first 500 principal components as features, which explained about 90% of the total variance in brain images and enabled model training in a reasonable period of time with advanced computational resources.

### Machine learning algorithms

Age estimation models were built using the sparse Bayesian relevance vector machine (RVM) in MATLAB (The MathWorks Inc, Natick, MA, USA),^24^ and the extreme gradient boosting package ‘xgboost’ v.0.82.1 in R.^25,94^ The RVM was developed as a probabilistic Bayesian equivalent to the popular support vector regression (SVR), and has widely been used in brain age research.^95–97^ We used the RVM implemented in MATLAB toolbox SparseBayes v.2 with the wrapper and kernel function by Qiu.^98^ Furthermore, we made use of the XGBoost algorithms, which have become popular methods after winning several machine learning challenges hosted by the data science competition platform Kaggle.^25,99^ XGBoost has previously been employed in brain age research.^15^ We used XGBoost with both the decision tree (‘gbtree’) and linear gradient booster (‘gblinear’). The learning rate was set to η = 0.02 with 5000 training iterations and an early stopping after 50 iterations in the case of no further model improvement. The maximum tree depth was set to 3. Default settings were used for all other training parameters.

### Model training and age prediction

Age estimation models were trained with the brain image PCA scores serving as features and chronological age serving as outcome variable. Model training and application was carried out in a 10-fold cross-validation manner with 100 repeats. Therefore, we randomly split the discovery sample into ten equal-sized subsets, of which nine subsets served for model training, and the remaining subset, the test sample, served for applying the model. Brain images of the training sample underwent PCA, and transformation parameters were subsequently applied to calculate PCA scores in the test sample. After the first model was trained and tested, the next subset served as test sample, while the other nine subsets were selected as training sample. This strategy was carried on until each subset served exactly once as test sample. The tenfold cross-validation procedure was repeated 100 times, so that 100 predictions were made for each subject. This procedure was performed for each tissue (grey and white matter) and model type (relevance vector machine, xgboost tree, and xgboost linear), resulting in a total number of 600 brain-predicted age estimates per subject. In a nested 10-fold cross-validation approach, we stacked the estimates from the three different model types in an ensemble estimate, resulting in 100 brain-predicted age estimates for grey matter, white matter, and combined grey and white matter, respectively. Finally, these estimates were averaged, leaving one brain-predicted age estimate for grey matter, white matter, and combined grey and white matter for each subject.

In the replication samples, we employed all discovery models from the 10-fold cross-validation procedure and compared the resulting age predictions against those derived from additional models where the complete discovery sample was used for model training. Results were highly concordant for all three tissue classes (*r* > .997). Due to the improved practicability, we performed subsequent replication analyses based on models trained on the complete discovery sample, which revealed virtually identical age predictions.

### Cross-trait association analysis

Cross-trait association analyses were carried out using PHESANT,^29^ an automated preprocessing and analysis pipeline for phenome-wide association analyses in UK Biobank datasets. Cross-trait associations were conducted for each BAG trait and 7,088 non-imaging derived UK Biobank variables. Sex, age, age^2^, scanner site, and total intracranial volume served as covariates. Based on variable type (continuous, integer, categorical single choice, or categorical multiple choice) and number of distinct values observed, different types of regression analyses (linear, logistic, ordinal logistic, or multinomial logistic) were performed. Variables suitable for linear regression underwent inverse-rank normal transformation. To obtain standardized effect size estimates, we converted the resulting beta coefficients from the different types of regression models to a corresponding correlation coefficient *r* based on *p*-value, number of observations, and number of covariates. Phenome-wide association analysis plots were created by assigning each variable to a custom UK Biobank category based on the respective variable’s UK Biobank data dictionary path.

### FreeSurfer associations

In addition to PheWAS analyses of non-imaging-derived phenotypes, we associated BAG with brain measures derived from the FreeSurfer aparc and aseg output files.^30^ FreeSurfer is an open-source software package to process, analyze and visualize human brain MR images. We retrieved FreeSurfer output files from the UKB resource (data-field 20263) and extracted surface area, cortical thickness, and cortical volume estimates of 34 bilateral cortical segmentations, as well as volume estimates of 8 bilateral subcortical segmentations, resulting in 220 brain measures in total. We calculated partial product-moment correlations between BAG and the 220 brain measures using sex, age, age^2^, scanner site, and total intracranial volume as covariates. Visualizations were performed using the ENIGMA toolbox.^100^

### UKB genotyping and imputation

We retrieved called and imputed genotypes (version 3) in BED and BGEN format, respectively, from the UK Biobank resource. Genotype collection, processing, and quality control have previously been described in detail.^23,101^ In brief, DNA was extracted from EDTA-treated whole-blood samples, aliquoted across three tubes (primary storage, backup storage, and genotyping tube), and shipped on 96-well plates of 50 µL aliquot per sample for genotyping to the Affymetrix Research Services Lab, Santa Clara, CA, USA. Genotyping was carried out using two arrays with a 95% marker overlap: the Applied Biosystems UK BiLEVE Axiom Array (807,411 markers; used in 49,950 participants) and the Applied Biosystems UK Biobank Axiom Array (825,927 markers; used in 438,427 participants). Both genotyping arrays were designed for genome-wide coverage of genetic content including biallelic single nucleotide polymorphisms (SNPs) and short insertions and deletions (indels). Marker-based quality control included testing for batch, plate, array, and sex effects, departures from Hardy-Weinberg-Equilibrium, as well as discordance across two control DNA replicates from the 1000 Genomes project, with two wells on each plate assigned to these control subjects. Sample-based quality control included missing rates (> 0.05), unusually high fractions of heterozygous variant calls, and sex chromosome aneuploidy. Relatedness between individuals was inferred from kinship coefficients estimated using KING.^102^ Population stratification was measured by applying fastPCA^103^ Principal Component Analysis on a set of 147,604 pruned high-quality markers. White-British ancestry (data-field 22006) was derived from a combination of self-report and genetic principal components. Genotype calls were phased using SHAPEIT3 and imputation was done using IMPUTE4 (https://jmarchini.org/software/) with the Haplotype Reference Consortium, UK10K, and 1000 Genomes phase 3 datasets serving as reference panels. Imputation resulted in ∼97M markers available for downstream analyses.

### LIFE-Adult genotyping and imputation

Genotype collection, processing, and quality control in LIFE-Adult have previously been described in detail.^104^ DNA was extracted from peripheral blood leukocytes. Genotyping was carried out using the Applied Biosystems Axiom Genome-Wide CEU 1 Array Plate with 587,352 markers. Marker-based quality control included call rate < 0.97, Hard-Weinberg equilibrium *p* < 1.0e-06, and plate association *p* < 1.0e-07. Sample quality control included dish-QC < 0.82, missing rates > 0.03, reported vs. genetic sex mismatch, and cryptic relatedness. Genotypes were pre-phased using SHAPEIT. Imputation was carried out using IMPUTE2 with the 1000 genome phase 3 dataset serving as reference. This resulted in 85,063,807 markers derived from 7,776 individuals. Post-imputation quality control included MAF ≥ 0.01 and INFO ≥ 0.8. As LIFE-Adult replication results were aggregated meta-analytically with the UKB replication results, we only included variants identified as biallelic SNPs or indels with INFO ≥ 0.8 in the UKB dataset (see ‘UKB genotyping and imputation’). This resulted in 9,472,504 markers passing quality-control in LIFE-Adult.

### Control for population structure

In the discovery sample, we calculated 20 genetic principal components by applying the randomized PCA algorithm (--pca 20 approx) implemented in PLINK v2.00a2LM 64-bit Intel (31 Jul 2019)^105^ on the same variants used by the UKB group (resource 1955; 146,988 markers passing our own quality-checks). For the UKB replication samples, we used genetic principal components provided by the pan-ancestry UKB project (UKB return 2442). The number of principal components serving as covariates was adjusted to the respective replication sample size. We used 10 principal components as covariates in the larger European-ancestry UKB replication sample and 3 principal components in each of the other UKB replication samples. In LIFE-Adult, 4 genetic principal components were used to account for subpopulation structure.

### Heritability and partitioned heritability

Estimates of SNP-based heritability (*h*^2^_SNP_) were derived by applying the GCTA genomic-restricted maximum likelihood (GREML) algorithm to genetic relatedness matrices (GRMs).^32,106^ GRMs were calculated based on biallelic autosomal variants with MAF ≥ 0.01 and INFO ≥ 0.80. GREML analyses were run in the discovery sample, with sex, age, age^2^, genotyping array, scanner site, total intracranial volume, and the first 20 genetic principal component serving as covariates. We conducted LD score regression^31,79^ on the GWAS summary statistics to corroborate GREML heritability estimates. We used precalculated LD scores and regression weights from 10000 Genomes phase 3 European ancestry samples (eur_w_ld_chr.tar.bz2). We retained variants with MAF > 0.01 included in the HapMap3 panel after removal of the MHC region (w_hm3.noMHC.snplist.zip). In order to partition heritability by functional annotation, we conducted stratified LD score regression^107^ using the baseline-LD model v.2.2 with the 1000 genomes phase 3 regression weights and allele frequencies excluding the MHC region. We considered functional annotations reported among the 33 ‘main annotations’ by Gazal and colleagues.^108^ Annotations with FDR < 0.05 were regarded as significant after multiple testing correction.^109^

### Genome-wide association analysis and replication meta-analysis

GWAS analysis were run in PLINK v2.00a2LM (31 Jul 2019) based on allelic dosage data. We included autosomal (chr1-22), gonosomal (chrX and chrY), pseudoautosomal (chrXY), and mitochondrial variations (chrMT). Analyses were run with male and female dosage data on a 0-2 scale on diploid chromosomes (chr1-22, chrXY), 0-1 scale on regular haploid chromosomes (chrY and chrMT), and 0-2 scale on chrX. We selected biallelic SNPs and INDELs with MAF > 0.01 and INFO > 0.80. Biallelic variations were defined as variations without duplicate chromosomal coordinates and duplicate identifiers in the imputed variant files. We modelled additive genetic effects and used sex, age, age^2^, total intra-cranial volume, scanner site, type of genotyping array, and the first 20 genetic principal components as covariates (3-10 genetic principal components were used in the replication GWASs; see section ‘control of population structure’). In total, there were 9,669,404 markers available for the discovery GWAS in *n* = 32,634 white-British ancestry individuals. The number of markers passing quality-control in the replication GWASs ranged between 8,364,077 (East-Asian ancestry) and 15,302,441 (African ancestry). Results of the ancestry-stratified replication GWASs were aggregated by performing a fixed-effects inverse-variance-weighted meta-analysis in METAL.^110^ Variants with an aggregated *n* lower than 67% of the 90th quantile of all observed *n* (adapted from LDSC)^31^ and heterogeneity *p* < 1.0e-06 were discarded. This yielded replication meta-analysis results from 9,496,239 (grey matter), 9,496,243 (white matter), and 9,496,192 (grey and white matter) variants in up to *n* = 7,259 individuals.

### Identification of independent discoveries

In order to identify independently associated variations, we performed stepwise conditional analyses employing the COJO module in GCTA.^32,33^ We used a 10,000 kb window-size (--cojo-wind 10000), a collinearity cutoff of 0.9 (–cojo-collinear 0.9) and included variants reaching at least suggestive evidence in the discovery GWAS (--cojo-p 1e-6). We only considered multiple signals within one locus to be independent if the *p*-value of the subsidiary association signal did not increase by more than two orders of magnitudes. Variants were regarded as independent genome-wide significant discoveries if they reached *p* < 5.0e-08 in conditional analysis. All other variants with conditional *p* < 1.0e-06 were regarded as suggestive signals, which were not considered for fine mapping but tested for result consistency in replication analyses. We denote variants discovered through conditional analyses as ‘index variants’ (i.e., the top variant of an association signal). To identify independent discoveries across the three BAG GWAS, we selected all index variants identified through trait-wise conditional analyses and clumped these variants according to chromosomal position and linkage disequilibrium using PLINK v1.90b6.8 (--clump-r2 0.1 --clump-kb 10000).

### Definition of variant replication and power calculations

For replication analyses, we selected index variations from genome-wide significant loci (*p* < 5.0e-08), and index variations from another 45 suggestive loci (conditional *p*-values ranging from 1.0e-06 to 5.0e-08). Consistency between discovery and replication results were tested via sign tests, i.e., binomial tests based on the number of observations where replication effect directions agree with the corresponding discovery effect directions. Variants with replication *p* < 0.05 (one-tailed nominal significance) were regarded as replicated variants. To estimate the expected number of successful replications, we carried out power calculations based on standardized beta coefficients (discovery), MAF (discovery), and N (replication).^111,112^ Beta coefficients were corrected for winner’s curse.^113^ The expected number of successful replications was calculated as sum of all power values obtained for each individual variant.

### Novelty of the discovered loci

Novelty of the discovered loci was examined by comparing our results against four previous genetic studies on BAG reporting discoveries at genome-wide significance levels.^17,18,20,21^ We used the clumping algorithm in PLINK v1.90b6.8 and regarded our own discoveries as novel if they did not clump together with previously reported variants, using a linkage disequilibrium threshold of *R*^2^ = 0.1 and a clumping window of 10 Mbp (--clump-r2 0.1 --clump-kb 10000).

### ANNOVAR enrichment test

We used the ANNOVAR (2017-07-17)^35^ enrichment test implemented in FUMA v.1.3.7, a web-based platform to functionally map and annotate GWAS results (https://fuma.ctglab.nl/),^114^ to test if genome-wide significant regions include relatively high or low proportions of variants with certain functional annotations. All candidate variants in linkage disequilibrium with an independent significant variant were considered for the ANNOVAR enrichment test. Independent significant variants were defined as autosomal variants reaching *p* < 5.E-08 and clumped with an *r*^2^ threshold of 0.60. Candidate variants were defined as all variants reaching *p* < 0.05 and showing *r*^2^ ≥ 0.60 with an independent significant variant. UK Biobank release2 served as reference panel. If variants were annotated with multiple functional categories, each category was counted as distinct annotation. Enrichment was computed as the proportion of variants with a certain annotation divided by the proportion of variants with that annotation relative to all available SNPs in the reference panel. A two-tailed Fisher’s exact test was conducted to test significance.

### Credible sets of variants

For each locus identified for the three BAG traits, we used FINEMAP^34^ v.1.4.1 to construct credible sets of variants that cumulatively capture 95% of the regional posterior probability to include the causal variant. FINEMAP uses a Bayesian framework and a computationally efficient shotgun stochastic search algorithm to model the LD structure and the strength of the variants’ associations (Z scores) to infer likely causal variants. For each locus, we used a 5 Mb window around the index variant to identify the furthest variants in linkage disequilibrium (*R*^2^ ≥ 0.1) with the index variation. Base pair positions of the identified variants were used as lower and upper bound of the respective genomic region. LD matrices covering all variants within that genomic region were calculated by applying LDstore^115^ v2.0 to the same allelic dosage data as employed for the genome-wide association analysis. We then applied FINEMAP allowing for up to *k*=10 causal variants within each region (‘--sss --n-causal-snps 10 --prob-cred-set 0.95’). Expected numbers of causal variants were derived by multiplying each evaluated number of *k* causal variants by the FINEMAP model-based probability. We report 95% credible sets for the most probable *k* model and report all variants that have been included in one of the credible sets for the *k* causal signals. We also report the FINEMAP model-averaged regional heritability, i.e., the estimated phenotypic variance explained by causal variants within each genomic region.^116^

### Functional annotation of variants

Annotation of variants was carried out using ANNOVAR,^35^ which allows to assign functional categories to variants through their physical position relative to defined genes. We used RefSeq gene annotations in human genome build 19 (hg19)^117^ retrieved from the UCSC Genome Browser Annotation Database.^118^ We identified the nearest gene based on the priority of the variant function (default ANNOVAR precedencies used) and the physical distance between the respective variant and the gene. Moreover, we used ANNOVAR to predict transcript consequences of non-synonymous exonic variants and added deleteriousness scores from CADD (Combined Annotation Dependent Depletion),^36^ DANN (Deep Neural Network for Annotating pathogenicity),^37^ and REVEL (Rare Exome Variant Ensemble Learner)^38^ provided in dataset dbnsfp35a (hg19).^119^ We also added information on the cytogenetic band of each variant.

### Gene nomination through functional annotation of credible variants

Credible variants were annotated using ANNOVAR (see section above),^35^ and variant posterior probabilities (see section ‘credible sets of variants’) were aggregated for each implicated gene. Genes were ranked according to their aggregated posterior probabilities and nominated for gene prioritization. Genes implicated by non-synonymous exonic variations were separately nominated for gene prioritization. Implicated genes were ranked based on the CADD phred-scaled scores of the non-synonymous exonic variant. If a gene was implicated by multiple non-synonymous variants, the top CADD phred-scaled score was used.

### Gene nomination through Summary-data-based Mendelian Randomization

We used summary-data-based Mendelian Randomization implemented in SMR v.1.03^40,41^ to test if the effect of an identified variant is potentially mediated by expression or splicing of a certain gene. The SMR software provides an integrative approach that combines GWAS summary statistics of complex phenotypes with information of omics resources to help prioritize gene targets and regulatory elements. It adopts the Mendelian Randomization (MR) strategy by using a genetic instrument (z) to test for pleiotropic association between gene regulation (exposure; x) and a trait of interest (outcome; y). The MR effect of gene regulation on a trait (*β*_xy_) is calculated as two-step least squares estimate and defined as the ratio of the instrument effect on the outcome (*β*_zy_) and that on the exposure (*β*_zx_), i.e., *β*_xy_ = *β*_zy_/*β*_yz_. The term ‘pleiotropy’ is preferred over ‘causality’ in this context, since SMR is based on a single genetic instrument and is unable to distinguish between (horizontal) pleiotropy and causality (vertical pleiotropy). The SMR software features the heterogeneity in dependent instruments (HEIDI) method, which uses multiple instruments in the regulatory region to distinguish pleiotropy from linkage (i.e., transcript and phenotype are not associated because of a shared causal variant but due to two or more distinct causal variants in linkage). We employed SMR with the BrainMeta v2 cis-eQTL (gene expression) and cis-sQTL (gene splicing) summary statistics derived from RNA-sequencing data of 2,865 brain cortex samples from 2,443 unrelated individuals of European ancestry.^42^ Our GWAS variants were succesfully mapped to 16,375 eQTL and 58,941 sQTL probes. We only considered results with FDR < 0.05, *p*_HEIDI_ > 0.01, and those associations that could be assigned to genome-wide significant loci from our discovery GWAS. We used the clumping procedure implemented in PLINK v1.90b6.8 to assign significant SMR associations to discovered index variations with a window-size of 3,000 kbp and *R*^2^ of 0.80. Genes implicated by eQTL and sQTL SMR were separately nominated for gene prioritization and ranked according to SMR p-value.

### Gene nomination through GTEx eQTL lookup

Index variations and their genome-wide significant neighbors in strong linkage disequilibrium (R^2^ > 0.8) were mapped to single-tissue and multi-tissue cis-QTLs cataloged in the Genotype-Tissue Expression (GTEx) V8 database.^120^ Significant variant-gene pairs for 49 tissues were obtained using the prefiltered file provided by GTEx (GTEx_Analysis_v8_eQTL.tar). Multi-tissue QTLs were obtained using the METASOFT results for all 49 tissues (GTEx_Analysis_v8.metasoft.txt.gz). Significant multi-tissue cis-QTLs were defined as variant-gene associations with data available for at least 10 tissues and with an m-value ≥ 0.9 (i.e., the posterior probability that the effect exists) in at least 50% of the available tissues. Of the remaining multi-tissue QTLs, a small fraction did not show convincing meta-analytic *p*-values (across tissues) derived from the Han and Eskin’s Random Effects Model (RE2). We thus set an inclusion criterion of RE2 *p* < 5E-8 (met by 99.9% of remaining entries), which resulted in a final number of 4,420,841 multi-tissue QTLs. Mapping our genome-wide significant variants to single- and multi-tissue QTLs was carried out using the GTEx hg19 liftover variant IDs. If multiple variants per discovered locus were associated with the expression of the same gene in the same tissue (single-tissue QTL mapping) or associated with the expression of the same gene across tissues (multi-tissue QTL mapping), respectively, we decided to only report QTL results of the variant that is in strongest LD to the index variation of the discovered locus.

We used the Bioconductor package biomaRt^121^ to convert Ensemble gene IDs to HGNC gene IDs and symbols. Genes implicated by GTEx single-tissue and multi-tissue eQTLs were separately nominated for gene prioritization and ranked according to the number of significant associations across tissues.

### Gene nomination through Polygenic Priority Scores

We calculated polygenic priority scores using PoPS,^43^ a similarity-based gene prioritization method designed to pinpoint causal genes in significant GWAS loci. PoPS is applied on gene-based results derived from MAGMA, and uses the full polygenic signal (including signals beneath genome-wide significance) to identify the most likely causal genes. PoPS incorporates data from a variety of sources, including 73 publicly available single-cell RNA sequencing datasets, curated biological pathways, and protein-protein interaction (PPI) networks. In total, more than 57,000 features are used to prioritize genes. We used the same PoPS feature map and same MAGMA gene annotation file as employed in the original work (downloaded from https://www.finucanelab.org/data).^43^ We applied MAGMA v.1.10 on our GWAS summary statistics using the SNP-wise mean gene analysis with linkage information derived from the discovery dataset (32,634 white-British ancestry individuals). For each index variant identified through conditional analyses, we nominated up to three genes with the highest PoPS scores for gene prioritization. Only genes located within 500 kb of the index variant and showing positive scores were considered.

### Gene prioritization

Genes considered for gene prioritization were nominated based on seven categories: 1) genes implicated by functional annotation of credible variants (ranked by cumulative variant posterior probabilities), 2) genes implicated by transcript consequences of non-synonymous exonic credible variants (ranked by highest CADD deleteriousness score), 3) genes implicated by SMR eQTLs (ranked by p-value), 4) genes implicated by SMR sQTLs (ranked by p-value), 5) genes implicated by GTEx single-tissue eQTLs (ranked by number of significant associations across tissues), 6) genes implicated by GTEx multi-tissue eQTLs (ranked by number of significant associations across tissues), 7) genes implicated by PoPS (ranked by score). For each nominated gene, we calculated a priority score as described below.

Let *i* represent a specific gene, and *j* denote the index of the gene nomination analysis conducted. The priority score (*P*_i_) for gene *i* is computed by considering both the cumulative posterior probability (*C*_i_) obtained from the first gene nomination category and the ranks of the gene (*R*_ij_) across the six additional nomination categories (*j* ∈ [1,6]). The formula is expressed as:

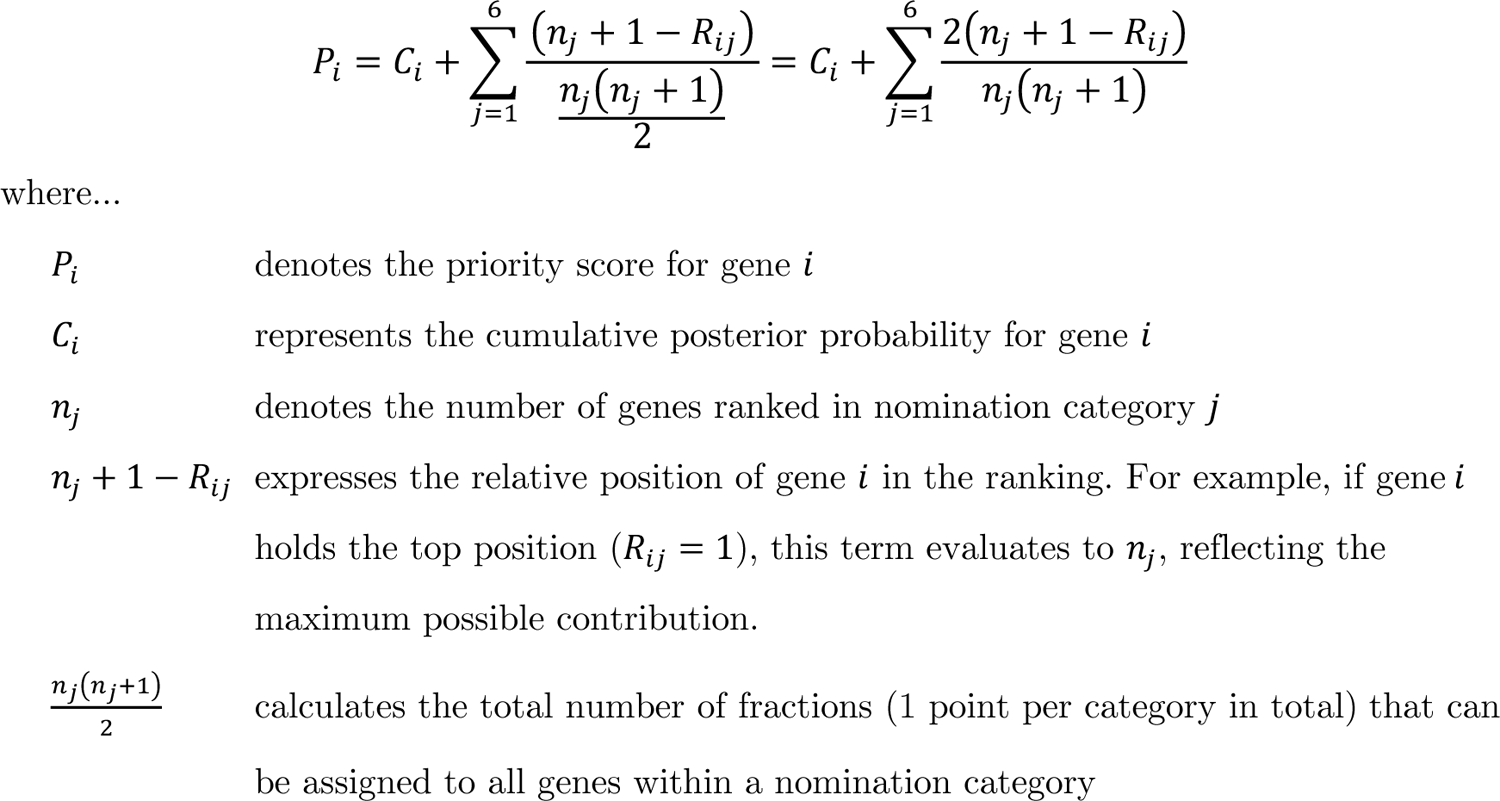

We designate the gene with the highest priority score per locus as the prioritized gene, indicating that it holds the highest probability of being causal.

### GWAS catalog search

All index variations from the SNP-level analyses and their genome-wide significant neighbors in strong linkage disequilibrium (R^2^ > 0.8) were selected for an NHGRI (National Human Genome Research Institute) GWAS catalog search. Index variations were identified through conditional analysis (see section ‘Conditional analysis’). Genome-wide significant neighbors in strong linkage disequilibrium were identified by carrying out a *p*-value informed clumping procedure with R2 > 0.8 and 3,000 kb window-size implemented in PLINK v1.90b6.8. We used the GWAS catalog released on December 21, 2021 (gwas_catalog_v1.0-associations_e105_r2021-12-21.tsv received from https://www.ebi.ac.uk/gwas/api/search/downloads/full). We only considered GWAS catalog entries that met genome-wide significance.

### Gene-based analysis

Gene-based analysis were carried out using fastBAT as implemented in GCTA 1.93.1f.^69^ Gene coordinates were obtained from the RefSeq gene annotation file in GFF3 format (genome-build GRCh37.p13; annotation release 105.20201022).^117^ NCBI chromosome names were converted to UCSC format. We selected genes of biotype ‘protein_coding’ located on chromosomes 1-22, X, Y, and MT. In the case of duplicate gene names, only the first entry was kept after sorting by chromosome, gene symbol, start coordinate, and end coordinate. This resulted in 19,319 protein-coding genes, of which 18,583 were successfully mapped to SNPs and INDELs included in GWAS analyses. We ran fastBAT with linkage information derived from the discovery dataset (32,634 white-British ancestry individuals). The window around gene boundaries was set to 0 kb to reduce dependencies between gene associations. Genes with FDR < 0.05 (Benjamini & Hochberg) were regarded as significant after multiple testing correction. To identify independent discoveries, we carried out a *p*-value informed clumping procedure with a conservative window-size of 3,000 kbp. To identify distinct discoveries across the three BAG traits, we performed a 2nd-level clumping procedure based on the top *p*-value of each gene across traits, again with 3,000 kbp window-size.

### Pathway analyses

Gene Ontology (GO) pathway analyses were conducted using R package GOfuncR 1.14.0 with Bioconductor database Homo.sapiens v1.3.1 build upon GO.db v3.14.0.^71,72,122,123^ The GO annotation knowledgebase is a curated collection of biological terms and their relationships in order to categorize genes and their products based on their involvement in ‘molecular processes’, the ‘cellular components’ where they perform actions, and the higher-order ‘biological processes’ they contribute to. We used a set of selected genes (genes prioritized through positional and transcription-based colocalization strategies and those with gene-based FDR < 0.05) to perform overrepresentation analyses of GO terms using the hypergeometric test implemented in GOfuncR. The total count of genes included in overrepresentation tests was 201, 339, and 303 for grey matter, white matter, and combined grey and white matter brain age, respectively. We also conducted gene set enrichment analyses (GSEA) based on the complete gene-based result table to test for lower-than-expected ranks (p-values) of gene sets using the Wilcoxon rank-sum test implemented in GOfuncR. By default, GOfuncR calculates family-wise error rates for terms in each of the three GO aspects based on random permutations. To reduce the chance of false discoveries, we integrated these permutation-based results to calculate family-wise error rates across the three GO aspects. In addition, we refined results with FWER < 0.05 by decorrelating GO terms and restricting results to the most specific terms using the implemented elim algorithm.^124^ For result evaluation and interpretation, we determined the number of distinct loci of genes that contribute to a GO term (using a 3,000 kbp clumping window) in order to account for spatial clustering of genes and potential gene result dependencies.

### Polygenic score analysis in replication sample

We used PRSice-2 to calculate polygenic scores (PGS) for BAG in the well-powered UKB European ancestry replication sample.^125^ Polygenic scores were computed based on variants reaching ten pre-defined discovery *p* thresholds: 1.00, 0.50, 0.20, 0.10, 0.05, 0.01, 1.0e-03, 1.0e-04, 1.0e-06, 5.0e-08.^82,126^ Variants with replication MAF < 0.01 were discarded. Variants were clumped with R^2^ < 0.1 and 500 kbp window-size. Associations between the ten resulting PGS and BAG were calculated as partial correlations using sex, age, age^2^, scanner site, total intracranial volume, genotyping array, and the first 10 genetic principal components serving as covariates.

### Genetic correlations

We carried out bivariate LD score regression^79^ to compute pair-wise genetic correlations both among BAG traits and between BAG traits and other complex traits. Bivariate LD score regression was run with 38 selected traits, which have frequently been used in similar investigations and cover a broad range of mental and physical health domains.^80–82^ In addition, we calculated genetic correlations with a set of 989 heritable UK biobank traits, whose GWAS summary statistics have been made publicly available by Neale and colleagues (Zenodo: https://doi.org/10.5281/zenodo.7186871). LD score regression analyses included HapMap3 variants after exclusion of variants in the MHC region (variant list downloaded from http://ldsc.broadinstitute.org/static/media/w_hm3.noMHC.snplist.zip). Genetic correlations with FDR < 0.05 were regarded as significant after multiple testing correction.

### Mendelian randomization

Evidence for potential causal associations between BAG and other complex traits was derived using generalized summary-data-based Mendelian Randomization (GSMR) as implemented in GCTA 1.93.1f.^32,83^ The GSMR method uses multiple genetic variants as instruments (z) to test for causality between an exposure (x) and outcome variable (y). Instruments are near-independent genetic variants (clumped with R^2^ < 0.05) associated with the exposure variable at a genome-wide significance level. GSMR is designed for two-sample scenarios, that is, GWAS summary-statistics of large independent studies are used to estimate the effects of the exposure on the outcome (*β*_xy_) based on the effects of the instruments on the exposure (*β*_zx_) and the effects of the instruments on the outcome (*β*_zy_). The ratio between *β*_xy_ and *β*_zx_ reveals the estimated mediation effect, i.e., *β*_xy_ = *β*_zy_/*β*_zx_, which is expected to be identical at each instrument under a causal model. Estimates from multiple instruments are integrated in a generalized least-squares approach. The use of multiple instruments enables to distinguish between causality, where the effect of an instrument on the outcome is mediated by the exposure, and horizontal pleiotropy, where the effect of an instrument on the outcome is exhibited through pathways other than the exposure. GSMR tests for heterogeneity in dependent instruments (HEIDI) to remove outliers based on the deviation of each instrument from the causal model. We used the default setting of removing HEIDI-outliers with deviation *p* < 0.01. To facilitate effect size comparisons, we standardized instrument effects on continuous exposure variables (*β*_zx_) based on z-statistic, allele frequency and sample size. GSMR has been demonstrated in simulations to be more powerful than inverse-variance-weighted MR (MR-IVW) and Egger regression (MR-Egger).^83^ Other empirical investigations have revealed qualitatively similar results between GSMR, MR-IVW, and MR-Egger.^82^ For GSMR analyses, we selected 11 risk factors based on the availability of summary statistics from large-scale GWAS that did not include the UK Biobank cohort and provided at least *m* = 30 independent genome-wide significant variants as instruments (clumped with *R*^2^ < 0.05). Due to the limited number of genome-wide significant variants for BAG, we only conducted unidirectional GSMR analyses with BAG serving as outcome variable.

### Polygenicity

We conducted genetic effect-size distribution inferences implemented in R package GENESIS v1.0^127^ to estimate the number of underlying susceptibility variants for BAG under a normal-mixture model of variant effects. Analyses of the benchmark traits neuroticism and height were based on the publicly available GWAS summary statistics by Baselmans et al.^87^ and Allen et al.^128^ Variants were filtered to 1.07 million common variants with MAF ≥ 0.05 included in the HapMap3 panel after exclusion of the major histocompatibility complex (MHC) region. SNPs with less than 0.67 times the 90th percentile of sample sizes and those with extremely large effect sizes (z^2^ > 80) were removed. We fitted the GENESIS three-component model, which assumes that 99% of the variant effects are null and the remaining 1% follow a mixture of two normal distributions, allowing a fraction of the susceptibility SNPs to belong to a cluster with larger effect sizes. We have chosen the three-component model (M3) over the simpler two-component model (M2), because a) M3 has been shown to provide distinctly better fits for a variety of complex traits, b) M3 has been shown to perform well even if the true data does not conform the model assumptions, and c) M2 appears to exhibit a more pronounced bias towards underestimating the number of susceptibility SNPs, particularly if model assumptions are not met.^89^ We used default settings for defining tagging SNPs and LD scores by using an LD cutoff of 0.1 and LD window of 1 Mbp.

## References

1. Kirkwood, T. B. L. Understanding the odd science of aging. Cell vol. 120 437–447 (2005).

2. da Silva, R., Conde, D. A., Baudisch, A. & Colchero, F. Slow and negligible senescence among testudines challenges evolutionary theories of senescence. Science (80-.). 376, 1466–1470 (2022).

3. Finch, C. E. & Austad, S. N. History and prospects: symposium on organisms with slow aging. Exp. Gerontol. 36, 593–597 (2001).

4. Vos, T. et al. Years lived with disability (YLDs) for 1160 sequelae of 289 diseases and injuries 1990&#x2013;2010: a systematic analysis for the Global Burden of Disease Study 2010. Lancet 380, 2163–2196 (2012).

5. Franke, K. & Gaser, C. 10 years of BrainAGE as an neuroimaging biomarker of brain aging: What insights did we gain? Front. Neurol. 10, 789 (2019).

6. Cole, J. H., Marioni, R. E., Harris, S. E. & Deary, I. J. Brain age and other bodily ‘ages’: implications for neuropsychiatry. Mol. Psychiatry 24, 266–281 (2019).

7. Liem, F. et al. Predicting brain-age from multimodal imaging data captures cognitive impairment. Neuroimage 148, 179–188 (2017).

8. Franke, K., Gaser, C., Manor, B. & Novak, V. Advanced BrainAGE in older adults with type 2 diabetes mellitus. Front. Aging Neurosci. 5, 90 (2013).

9. Franke, K., Ristow, M. & Gaser, C. Gender-specific impact of personal health parameters on individual brain aging in cognitively unimpaired elderly subjects. Front. Aging Neurosci. 6, (2014).

10. Steffener, J. et al. Differences between chronological and brain age are related to education and self-reported physical activity. Neurobiol. Aging 40, (2016).

11. Richard, G. et al. Assessing distinct patterns of cognitive aging using tissue-specific brain age prediction based on diffusion tensor imaging and brain morphometry. PeerJ 2018, (2018).

12. Cole, J. H. et al. Brain age predicts mortality. Mol. Psychiatry 23, 1385–1392 (2018).

13. Jawinski, P. et al. Linking Brain Age Gap to Mental and Physical Health in the Berlin Aging Study II. Front. Aging Neurosci. 14, (2022).

14. Shahab, S. et al. Brain structure, cognition, and brain age in schizophrenia, bipolar disorder, and healthy controls. Neuropsychopharmacol. Off. Publ. Am. Coll. Neuropsychopharmacol. 44, 898–906 (2019).

15. Kaufmann, T. et al. Common brain disorders are associated with heritable patterns of apparent aging of the brain. Nat. Neurosci. 22, 1617–1623 (2019).

16. Cole, J. H. et al. Predicting brain age with deep learning from raw imaging data results in a reliable and heritable biomarker. Neuroimage 163, 115–124 (2017).

17. Jonsson, B. A. et al. Brain age prediction using deep learning uncovers associated sequence variants. Nat. Commun. 10, 5409 (2019).

18. Smith, S. M. et al. Brain aging comprises many modes of structural and functional change with distinct genetic and biophysical associations. Elife 9, e52677 (2020).

19. Ning, K. et al. Improving brain age estimates with deep learning leads to identification of novel genetic factors associated with brain aging. Neurobiol. Aging 105, 199–204 (2021).

20. Leonardsen, E. H. et al. Genetic architecture of brain age and its causal relations with brain and mental disorders. Mol. Psychiatry (2023) doi:10.1038/s41380-023-02087-y.

21. Ning, K., Zhao, L., Matloff, W., Sun, F. & Toga, A. W. Association of relative brain age with tobacco smoking, alcohol consumption, and genetic variants. Sci. Rep. 10, (2020).

22. Gaser, C., et al. CAT {\textendash} A Computational Anatomy Toolbox for the Analysis of Structural MRI Data. *bioRxiv* (2023) doi:10.1101/2022.06.11.495736.

23. Bycroft, C. et al. The UK Biobank resource with deep phenotyping and genomic data. Nature 562, 203–209 (2018).

24. Tipping, M. E. Sparse Bayesian Learning and the Relevance Vector Machine. J. Mach. Learn. Res. 1, 211–244 (2001).

25. Chen, T., et al. xgboost: Extreme Gradient Boosting. (2019).

26. Loeffler, M. et al. The LIFE-Adult-Study: objectives and design of a population-based cohort study with 10,000 deeply phenotyped adults in Germany. BMC Public Health 15, 691 (2015).

27. Engel, C. et al. Cohort Profile: The LIFE-Adult-Study. Int. J. Epidemiol. (2022) doi:10.1093/ije/dyac114.

28. McGraw, K. O. & Wong, S. P. Forming inferences about some intraclass correlation coefficients. Psychol. Methods 1, 30–46 (1996).

29. Millard, L. A. C., Davies, N. M., Gaunt, T. R., Davey Smith, G. & Tilling, K. Software Application Profile: PHESANT: a tool for performing automated phenome scans in UK Biobank. Int. J. Epidemiol. 47, 29–35 (2018).

30. Dale, A. M., Fischl, B. & Sereno, M. I. Cortical Surface-Based Analysis: I. Segmentation and Surface Reconstruction. Neuroimage 9, 179–194 (1999).

31. Bulik-Sullivan, B. et al. LD score regression distinguishes confounding from polygenicity in genome-wide association studies. Nat. Genet. 47, 291–295 (2015).

32. Yang, J., Lee, S. H., Goddard, M. E. & Visscher, P. M. GCTA: A tool for genome-wide complex trait analysis. Am. J. Hum. Genet. 88, 76–82 (2011).

33. Yang, J. et al. Conditional and joint multiple-SNP analysis of GWAS summary statistics identifies additional variants influencing complex traits. Nat. Genet. 44, 369– 75, S1-3 (2012).

34. Benner, C. et al. FINEMAP: efficient variable selection using summary data from genome-wide association studies. Bioinformatics 32, 1493–1501 (2016).

35. Wang, K., Li, M. & Hakonarson, H. ANNOVAR: functional annotation of genetic variants from high-throughput sequencing data. Nucleic Acids Res. 38, e164–e164 (2010).

36. Rentzsch, P., Schubach, M., Shendure, J. & Kircher, M. CADD-Splice—improving genome-wide variant effect prediction using deep learning-derived splice scores. Genome Med. 13, 31 (2021).

37. Quang, D., Chen, Y. & Xie, X. DANN: a deep learning approach for annotating the pathogenicity of genetic variants. Bioinformatics 31, 761–763 (2015).

38. Ioannidis, N. M. et al. REVEL: An Ensemble Method for Predicting the Pathogenicity of Rare Missense Variants. Am. J. Hum. Genet. 99, 877–885 (2016).

39. Ardlie, K. G. et al. The Genotype-Tissue Expression (GTEx) pilot analysis: Multitissue gene regulation in humans. Science (80-.). 348, 648–660 (2015).

40. Zhu, Z. et al. Integration of summary data from GWAS and eQTL studies predicts complex trait gene targets. Nat. Genet. 48, 481–487 (2016).

41. Wu, Y. et al. Integrative analysis of omics summary data reveals putative mechanisms underlying complex traits. Nat. Commun. 9, 918 (2018).

42. Qi, T. et al. Genetic control of RNA splicing and its distinct role in complex trait variation. Nat. Genet. (2022) doi:10.1038/s41588-022-01154-4.

43. Weeks, E. M. et al. Leveraging polygenic enrichments of gene features to predict genes underlying complex traits and diseases. Nat. Genet. 55, 1267–1276 (2023).

44. Stefansson, H. et al. A common inversion under selection in Europeans. Nat. Genet. 37, 129–137 (2005).

45. Okbay, A. et al. Genetic variants associated with subjective well-being, depressive symptoms, and neuroticism identified through genome-wide analyses. Nat. Genet. 48, 624–633 (2016).

46. Sollis, E. et al. The NHGRI-EBI GWAS Catalog: knowledgebase and deposition resource. Nucleic Acids Res. 51, D977–D985 (2023).

47. Lee, J. J. et al. Gene discovery and polygenic prediction from a genome-wide association study of educational attainment in 1.1 million individuals. Nat. Genet. 50, 1112–1121 (2018).

48. Nagel, M. et al. Meta-analysis of genome-wide association studies for neuroticism in 449,484 individuals identifies novel genetic loci and pathways. Nat. Genet. 50, 920–927 (2018).

49. Karlsson Linnér, R., et al. Genome-wide association analyses of risk tolerance and risky behaviors in over 1 million individuals identify hundreds of loci and shared genetic influences. Nat. Genet. 51, 245–257 (2019).

50. Jansen, P. R. et al. Genome-wide analysis of insomnia in 1,331,010 individuals identifies new risk loci and functional pathways. Nat. Genet. 51, 394–403 (2019).

51. Kichaev, G. et al. Leveraging Polygenic Functional Enrichment to Improve GWAS Power. Am. J. Hum. Genet. 104, 65–75 (2019).

52. Hollis, B. et al. Genomic analysis of male puberty timing highlights shared genetic basis with hair colour and lifespan. Nat. Commun. 11, 1536 (2020).

53. Jun, G. et al. A novel Alzheimer disease locus located near the gene encoding tau protein. Mol. Psychiatry 21, 108–117 (2016).

54. Rademakers, R., Cruts, M. & van Broeckhoven, C. The role of tau (MAPT) in frontotemporal dementia and related tauopathies. Hum. Mutat. 24, 277–295 (2004).

55. Bittner, S., Ruck, T., Fernández-Orth, J. & Meuth, S. G. TREK-King the Blood– Brain-Barrier. J. Neuroimmune Pharmacol. 9, 293–301 (2014).

56. Wang, W. et al. Lig4-4 selectively inhibits TREK-1 and plays potent neuroprotective roles in vitro and in rat MCAO model. Neurosci. Lett. 671, 93–98 (2018).

57. van der Meer, D. et al. Understanding the genetic determinants of the brain with MOSTest. Nat. Commun. 11, 3512 (2020).

58. Shadrin, A. A. et al. Vertex-wise multivariate genome-wide association study identifies 780 unique genetic loci associated with cortical morphology. Neuroimage 244, 118603 (2021).

59. Le Guen, Y. et al. eQTL of KCNK2 regionally influences the brain sulcal widening: evidence from 15,597 UK Biobank participants with neuroimaging data. Brain Struct. Funct. 224, 847–857 (2019).

60. Sharma, K., Tyagi, R., Singh, R., Sharma, S. K. & Anand, A. Serum Levels of TIMP-3, LIPC, IER3, and SLC16A8 in CFH-Negative AMD Cases. J. Cell. Biochem. 118, 2087–2095 (2017).

61. Zaharija, B., Samardžija, B. & Bradshaw, N. J. The TRIOBP Isoforms and Their Distinct Roles in Actin Stabilization, Deafness, Mental Illness, and Cancer. Molecules 25, (2020).

62. Vujkovic, M. et al. Discovery of 318 new risk loci for type 2 diabetes and related vascular outcomes among 1.4 million participants in a multi-ancestry meta-analysis. Nat. Genet. 52, 680–691 (2020).

63. Sakaue, S. et al. A cross-population atlas of genetic associations for 220 human phenotypes. Nat. Genet. 53, 1415–1424 (2021).

64. Malik, T. H. et al. Transcriptional repression and developmental functions of the atypical vertebrate GATA protein TRPS1. EMBO J. 20, 1715–1725 (2001).

65. Qi, T. et al. Identifying gene targets for brain-related traits using transcriptomic and methylomic data from blood. Nat. Commun. 9, 2282 (2018).

66. Cheung, C. T., Kaul, S. C. & Wadhwa, R. Molecular bridging of aging and cancer. Ann. N. Y. Acad. Sci. 1197, 129–133 (2010).

67. Fjorback, A. W., Müller, H. K. & Wiborg, O. Membrane glycoprotein M6B interacts with the human serotonin transporter. J. Mol. Neurosci. 37, 191–200 (2009).

68. Zhang, G. et al. DHRSX, a novel non-classical secretory protein associated with starvation induced autophagy. Int. J. Med. Sci. 11, 962–970 (2014).

69. Bakshi, A. et al. Fast set-based association analysis using summary data from GWAS identifies novel gene loci for human complex traits. Sci. Rep. 6, 1–9 (2016).

70. Jansen, I. E. et al. Genome-wide meta-analysis identifies new loci and functional pathways influencing Alzheimer’s disease risk. Nat. Genet. 51, 404–413 (2019).

71. Ashburner, M. et al. Gene Ontology: tool for the unification of biology. Nat. Genet. 25, 25–29 (2000).

72. Grote, S. GOfuncR: Gene ontology enrichment using FUNC. (2021).

73. Takai, Y., Sasaki, T. & Matozaki, T. Small GTP-binding proteins. Physiol. Rev. 81, 153–208 (2001).

74. Reiner, D. J. & Lundquist, E. A. Small GTPases. WormBook: the online review of C. elegans biology vol. 2018 1–65 (2018).

75. Wennerberg, K., Rossman, K. L. & Der, C. J. The Ras superfamily at a glance. J. Cell Sci. 118, 843–846 (2005).

76. Ejaz, A., Mattesich, M. & Zwerschke, W. Silencing of the small GTPase DIRAS3 induces cellular senescence in human white adipose stromal/progenitor cells. Aging (Albany. NY). 9, 860–879 (2017).

77. Wang, L., Yang, L., Debidda, M., Witte, D. & Zheng, Y. Cdc42 GTPase-activating protein deficiency promotes genomic instability and premature aging-like phenotypes. Proc. Natl. Acad. Sci. U. S. A. 104, 1248–1253 (2007).

78. Debidda, M., Williams, D. A. & Zheng, Y. Rac1 GTPase regulates cell genomic stability and senescence. J. Biol. Chem. 281, 38519–38528 (2006).

79. Bulik-Sullivan, B. et al. An atlas of genetic correlations across human diseases and traits. Nat. Genet. 47, 1236–1241 (2015).

80. Abdellaoui, A. & Verweij, K. J. H. Dissecting polygenic signals from genome-wide association studies on human behaviour. *Nat*. Hum. Behav. 5, 686–694 (2021).

81. Abdellaoui, A. et al. Genetic correlates of social stratification in Great Britain. *Nat*. Hum. Behav. 3, 1332–1342 (2019).

82. Wray, N. R. et al. Genome-wide association analyses identify 44 risk variants and refine the genetic architecture of major depression. Nat. Genet. 50, 668–681 (2018).

83. Zhu, Z. et al. Causal associations between risk factors and common diseases inferred from GWAS summary data. Nat. Commun. 9, 224 (2018).

84. Evangelou, E. et al. Genetic analysis of over 1 million people identifies 535 new loci associated with blood pressure traits. Nat. Genet. 50, 1412–1425 (2018).

85. Wu, W. et al. An electroencephalographic signature predicts antidepressant response in major depression. Nat. Biotechnol. 1–9 (2020) doi:10.1038/s41587-019-0397-3.

86. Wood, A. R. et al. Defining the role of common variation in the genomic and biological architecture of adult human height. Nat. Genet. 46, 1173–1186 (2014).

87. Baselmans, B. M. L. et al. Multivariate genome-wide analyses of the well-being spectrum. Nat. Genet. 51, 445–451 (2019).

88. Montag, C., Ebstein, R. P., Jawinski, P. & Markett, S. Molecular genetics in psychology and personality neuroscience: On candidate genes, genome wide scans, and new research strategies. Neuroscience and Biobehavioral Reviews vol. 118 163–174 (2020).

89. Zhang, Y., Qi, G., Park, J. H. & Chatterjee, N. Estimation of complex effect-size distributions using summary-level statistics from genome-wide association studies across 32 complex traits. Nat. Genet. 50, 1318–1326 (2018).

90. Gaser, C., Franke, K., Klöppel, S., Koutsouleris, N. & Sauer, H. BrainAGE in Mild Cognitive Impaired Patients: Predicting the Conversion to Alzheimer’s Disease. PLoS One 8, e67346 (2013).

91. Savage, J. E. et al. Genome-wide association meta-analysis in 269,867 individuals identifies new genetic and functional links to intelligence. Nat. Genet. 50, 912–919 (2018).

92. Vidal-Piñeiro, D. et al. Individual variations in “Brain age” relate to early life factors more than to longitudinal brain change. *bioRxiv* 2021.02.08.428915 (2021) doi:10.1101/2021.02.08.428915.

93. Frei, O. et al. Bivariate causal mixture model quantifies polygenic overlap between complex traits beyond genetic correlation. Nat. Commun. 10, 2417 (2019).

94. Larstone, R. M., Jang, K. L., Livesley, W. J., Vernon, P. A. & Wolf, H. The relationship between Eysenck’s P-E-N model of personality, the five-factor model of personality, and traits delineating personality dysfunction. Pers. Individ. Dif. 33, 25–37 (2002).

95. Franke, K., Ziegler, G., Klöppel, S. & Gaser, C. Estimating the age of healthy subjects from T1-weighted MRI scans using kernel methods: Exploring the influence of various parameters. Neuroimage 50, 883–892 (2010).

96. Franke, K., Luders, E., May, A., Wilke, M. & Gaser, C. Brain maturation: Predicting individual BrainAGE in children and adolescents using structural MRI. Neuroimage 63, 1305–1312 (2012).

97. Franke, K. & Gaser, C. Longitudinal changes in individual BrainAGE in healthy aging, mild cognitive impairment, and Alzheimer’s Disease. GeroPsych J. Gerontopsychology Geriatr. Psychiatry 25, 235–245 (2012).

98. Qiu, K. Relevance Vector Machine (RVM). (2019).

99. Chen, T. & Guestrin, C. XGBoost: A Scalable Tree Boosting System. in Proceedings of the 22nd ACM SIGKDD International Conference on Knowledge Discovery and Data Mining 785–794 (Association for Computing Machinery, 2016). doi:10.1145/2939672.2939785.

100. Larivière, S. et al. The ENIGMA Toolbox: multiscale neural contextualization of multisite neuroimaging datasets. Nat. Methods 18, 698–700 (2021).

101. Welsh, S., Peakman, T., Sheard, S. & Almond, R. Comparison of DNA quantification methodology used in the DNA extraction protocol for the UK Biobank cohort. BMC Genomics 18, 26 (2017).

102. Manichaikul, A. et al. Robust relationship inference in genome-wide association studies. Bioinformatics 26, 2867–2873 (2010).

103. Galinsky, K. J. et al. Fast principal-component analysis reveals convergent evolution of ADH1B in Europe and East Asia. Am. J. Hum. Genet. 98, 456–472 (2016).

104. Jawinski, P. et al. Human brain arousal in the resting state: a genome-wide association study. Mol. Psychiatry 24, 1599–1609 (2019).

105. Chang, C. C. et al. Second-generation PLINK: rising to the challenge of larger and richer datasets. Gigascience 4, s13742--015 (2015).

106. Yang, J. et al. Common SNPs explain a large proportion of the heritability for human height. Nat. Genet. 42, 565–569 (2010).

107. Finucane, H. K. et al. Partitioning heritability by functional annotation using genome-wide association summary statistics. Nat. Genet. 47, 1228–1235 (2015).

108. Gazal, S. et al. Functional architecture of low-frequency variants highlights strength of negative selection across coding and non-coding annotations. Nat. Genet. 50, 1600– 1607 (2018).

109. Benjamini, Y. & Hochberg, Y. Controlling the False Discovery Rate: A Practical and Powerful Approach to Multiple Testing. J. R. Stat. Soc. 57, 289–300 (1995).

110. Willer, C. J., Li, Y. & Abecasis, G. R. METAL: fast and efficient meta-analysis of genomewide association scans. Bioinformatics 26, 2190–2191 (2010).

111. Visscher, P. M. et al. 10 Years of GWAS Discovery: Biology, Function, and Translation. Am. J. Hum. Genet. 101, 5–22 (2017).

112. Adhikari, K. R Functions to calculate power of GWAS studies. https://github.com/kaustubhad/gwas-power (2018).

113. Palmer, C. & Pe’er, I. Statistical correction of the Winner’s Curse explains replication variability in quantitative trait genome-wide association studies. PLOS Genet. 13, e1006916 (2017).

114. Watanabe, K., Taskesen, E., van Bochoven, A. & Posthuma, D. Functional mapping and annotation of genetic associations with FUMA. Nat. Commun. 8, 1826 (2017).

115. Benner, C. et al. Prospects of Fine-Mapping Trait-Associated Genomic Regions by Using Summary Statistics from Genome-wide Association Studies. Am. J. Hum. Genet. 101, 539–551 (2017).

116. Benner, C., Havulinna, A. S., Salomaa, V., Ripatti, S. & Pirinen, M. Refining fine-mapping: effect sizes and regional heritability. *bioRxiv* (2018) doi:10.1101/318618.

117. O’Leary, N. A. et al. Reference sequence (RefSeq) database at NCBI: current status, taxonomic expansion, and functional annotation. Nucleic Acids Res. 44, D733–45 (2016).

118. Nassar, L. R. et al. The UCSC Genome Browser database: 2023 update. Nucleic Acids Res. 51, D1188–D1195 (2023).

119. Liu, X., Wu, C., Li, C. & Boerwinkle, E. dbNSFP v3.0: A One-Stop Database of Functional Predictions and Annotations for Human Nonsynonymous and Splice-Site SNVs. Hum. Mutat. 37, 235–241 (2016).

120. Aguet, F. et al. Genetic effects on gene expression across human tissues. Nature 550, 204–213 (2017).

121. Durinck, S., Spellman, P. T., Birney, E. & Huber, W. Mapping identifiers for the integration of genomic datasets with the R/Bioconductor package biomaRt. Nat. Protoc. 4, 1184–1191 (2009).

122. Team, B. C. Homo.sapiens: Annotation package for the Homo.sapiens object. (2015).

123. Carlson, M. GO.db: A set of annotation maps describing the entire Gene Ontology. (2021).

124. Alexa, A., Rahnenführer, J. & Lengauer, T. Improved scoring of functional groups from gene expression data by decorrelating GO graph structure. Bioinformatics 22, 1600– 1607 (2006).

125. Choi, S. W. & O’Reilly, P. F. PRSice-2: Polygenic Risk Score software for biobank-scale data. Gigascience 8, giz082 (2019).

126. Consortium, S. W. G. of the P. G. et al. Biological insights from 108 schizophrenia-associated genetic loci. Nature 511, 421 (2014).

127. Zhang, Y., Qi, G., Park, J. H. & Chatterjee, N. Estimation of complex effect-size distributions using summary-level statistics from genome-wide association studies across 32 complex traits. Nat. Genet. 50, 1318–1326 (2018).

128. Allen, H. L. et al. Hundreds of variants clustered in genomic loci and biological pathways affect human height. Nature 467, 832–838 (2010).

