## Supplementary Materials A (Figures) for "Genome-wide analysis of brain age identifies 25 associated loci and unveils relationships with mental and physical health"

### Supplementary Figures

#### Phenotypic results

Fig. A1 Prediction accuracies ........ 2
Fig. A2 Phenome-wide association analyses results ........ 3
Fig. A3 Phenotypic correlations with FreeSurfer measures ........ 4

#### Discovery genome-wide association analyses

Fig. A4 Enrichment of heritability stratified by functional annotation ........ 5
Fig. A5 Regional plots 1-6 for grey matter BAG ........ 6
Fig. A6 Regional plots 7-12 for grey matter BAG ........ 7
Fig. A7 Regional plots 1-6 for white matter BAG ........ 8
Fig. A8 Regional plots 7-12 for white matter BAG ........ 9
Fig. A9 Regional plots 13-16 for white matter BAG ........ 10
Fig. A10 Regional plots 1-6 for combined BAG ........ 11
Fig. A11 Regional plots 7-12 for combined BAG ........ 12
Fig. A12 Regional plot 13 for combined BAG ........ 13
Fig. A13 ANNOVAR enrichment test of functional consequences ........ 14

#### Replication genome-wide association analyses

Fig. A14 Manhattan and quantile-quantile-plots for grey matter BAG ........ 15
Fig. A15 Manhattan and quantile-quantile-plots for white matter BAG ........ 16
Fig. A16 Manhattan and quantile-quantile-plots for combined BAG ........ 17
Fig. A17 Replication GWAMAs: Manhattan and quantile-quantile q-plots ........ 18
Fig. A18 Quantile-quantile plots pf ........

#### Gene-based and Mendelian Randomization analyses

Fig. A19 Manhattan plots of gene-based association analyses ........ 19
Fig. A20 Mendelian Randomization results for grey matter BAG ........ 20
Fig. A21 Mendelian Randomization results for white matter BAG ........ 21
Fig. A22 Mendelian Randomization results for combined BAG ........ 22

**References** ........ 22

### Supplementary Figures

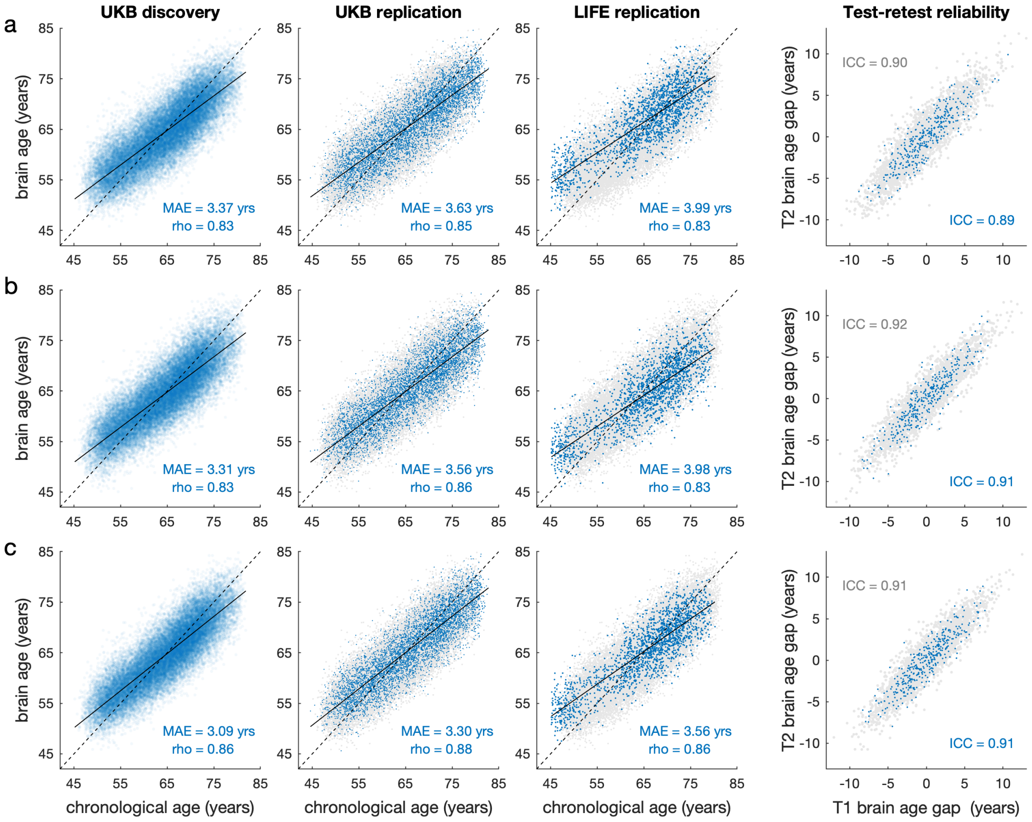


**Fig. A1** Accuracy of the age estimation models stratified by tissue class: **(a)** grey matter, **(b)** white matter, and **(c)** combined grey and white matter. Blue dots in the first three plots (from left to right) show brain-predicted age estimates plotted against the chronological age in the UKB discovery sample ($n$ = 32,634; white-British ancestry), UKB replication sample ($n$ = 5,426; cross-ancestry), and LIFE-Adult replication sample ($n$ = 1,883; European ancestry). To facilitate comparisons, results of the UKB discovery sample are also shown as grey dots in the background of the UKB replication and LIFE replication plots. At this stage, brain-predicted age estimates have not yet been bias-corrected for regression dilution, i.e., younger participants' ages are systematically overestimated and vice versa, as indicated by the linear regression line (solid) crossing the identity line (dashed). The fourth plot shows the test-retest reliabilities of brain age gap (i.e., the difference between brain-predicted and chronological age) in a subset of the UKB discovery (grey dots, $n$ = 3,625) and UKB replication sample (blue dots, $n$ = 376). For test-retest comparisons, brain age gap was bias-corrected for age, age$^2$, sex, scanner site, and total intracranial volume. MAE: mean absolute error; rho: product-moment correlation coefficient. ICC: intraclass correlation coefficient (C,1).(McGraw and Wong, 1996)

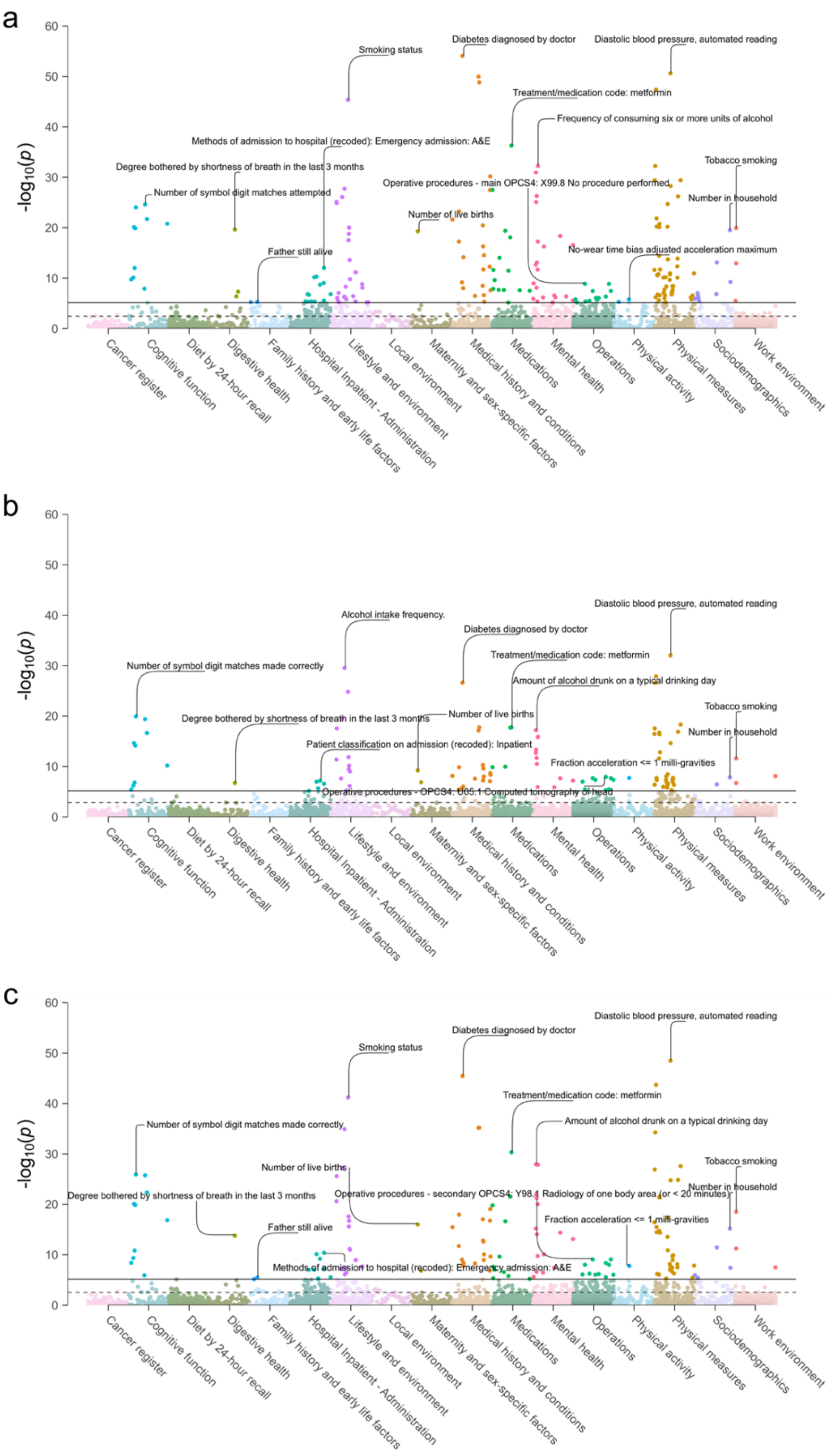


**Fig. A2** Phenome-wide association results: Each dot represents a single association between brain age gap and one out of 7,088 non-imaging derived UK Biobank phenotypes. P-values (-log10 scale) are shown on the y axis, with phenotypes on the x axis categorized according to their UK Biobank data dictionary path and ordered alphanumerically. Horizontal lines indicate the Bonferroni-adjusted (solid line) and FDR-adjusted (dashed line) level of significance. The top associations per category have been annotated. **(a)** grey matter brain age gap, **(b)** white matter brain age gap, **(c)** combined grey and white matter brain age

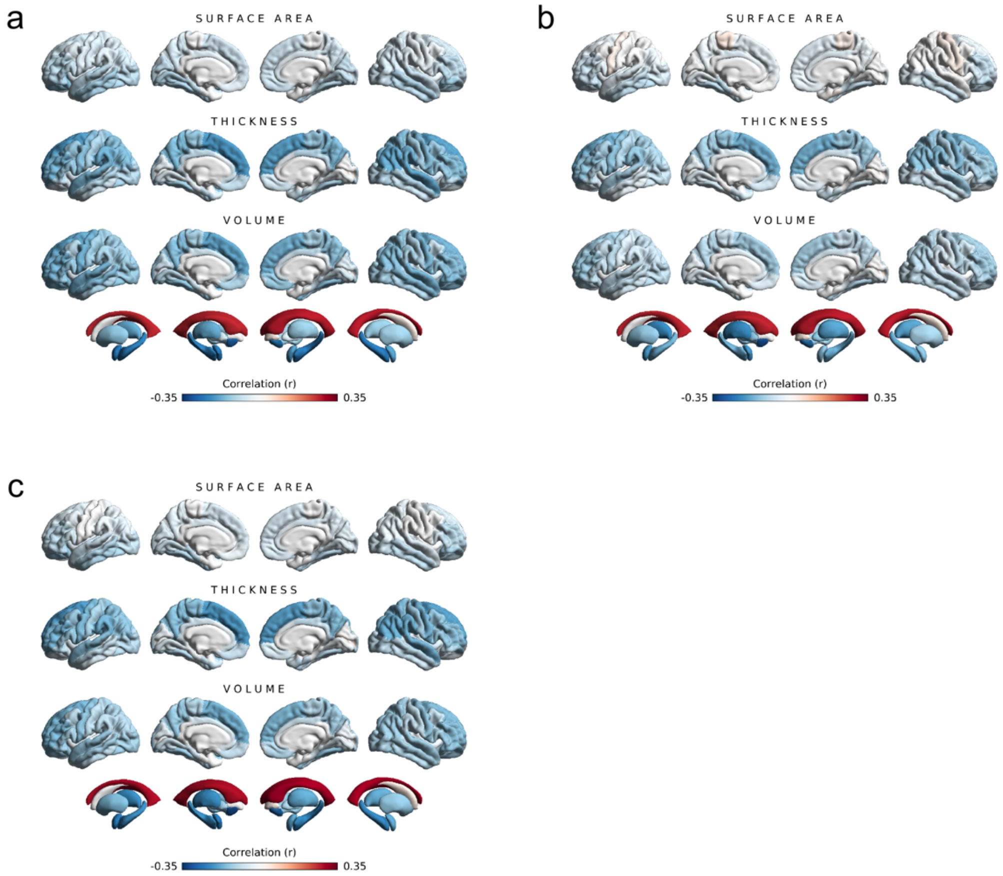


**Fig. A3** Surface plots showing the correlations between brain age gap and 220 brain structure variables obtained from the FreeSurfer aparc and aseg output (surface area, cortical thickness, and cortical volume of 34 bilateral cortical segmentations, as well as volume of 16 bilateral subcortical segmentations). Colors reflect the strength and direction of partial product-moment correlations (adjusted for sex, age, age2, assessment center, total intracranial volume). Plots have been created using the ENIGMA toolbox in MATLAB. **(a)** grey matter brain age gap, **(b)** white matter brain age gap, **(c)** combined grey and white matter brain age gap.

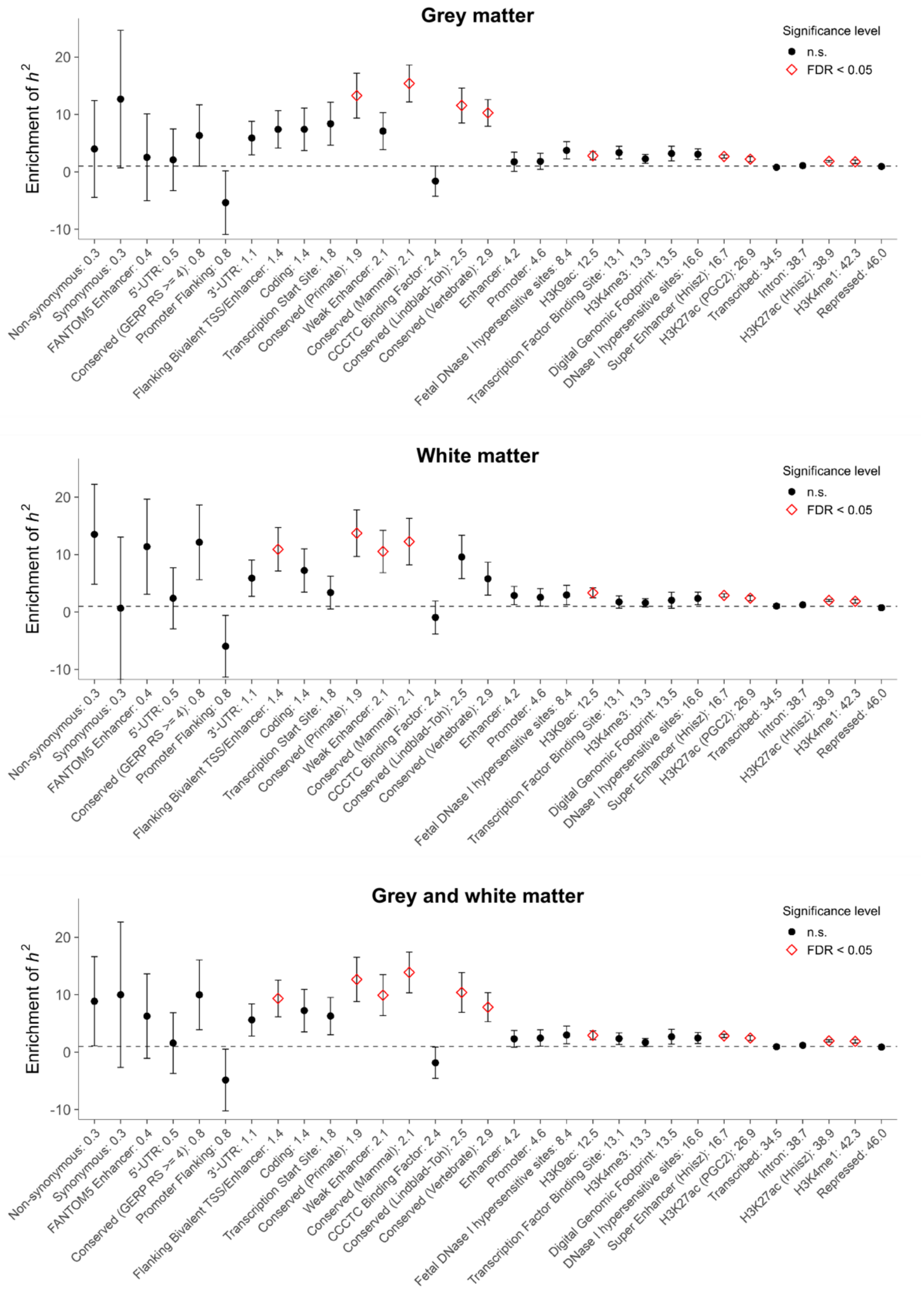


**Fig. A4** Enrichment of heritability ($h^2$) in genomic regions partitioned by functional category. Results refer to grey matter (top) white matter (middle) and combined grey and white matter brain age gap (bottom). Enrichment of $h^2$ of a category is defined as proportion of $h^2$ attributable to the category divided by the proportion of genetic variants in that category. Error bars represent jackknife standard errors around the estimates of enrichment. Functional annotations have been ordered by proportion of SNPs contained within the categories (from left to right; percentages shown at x axis ticks). Significant categories are highlighted as red diamonds.

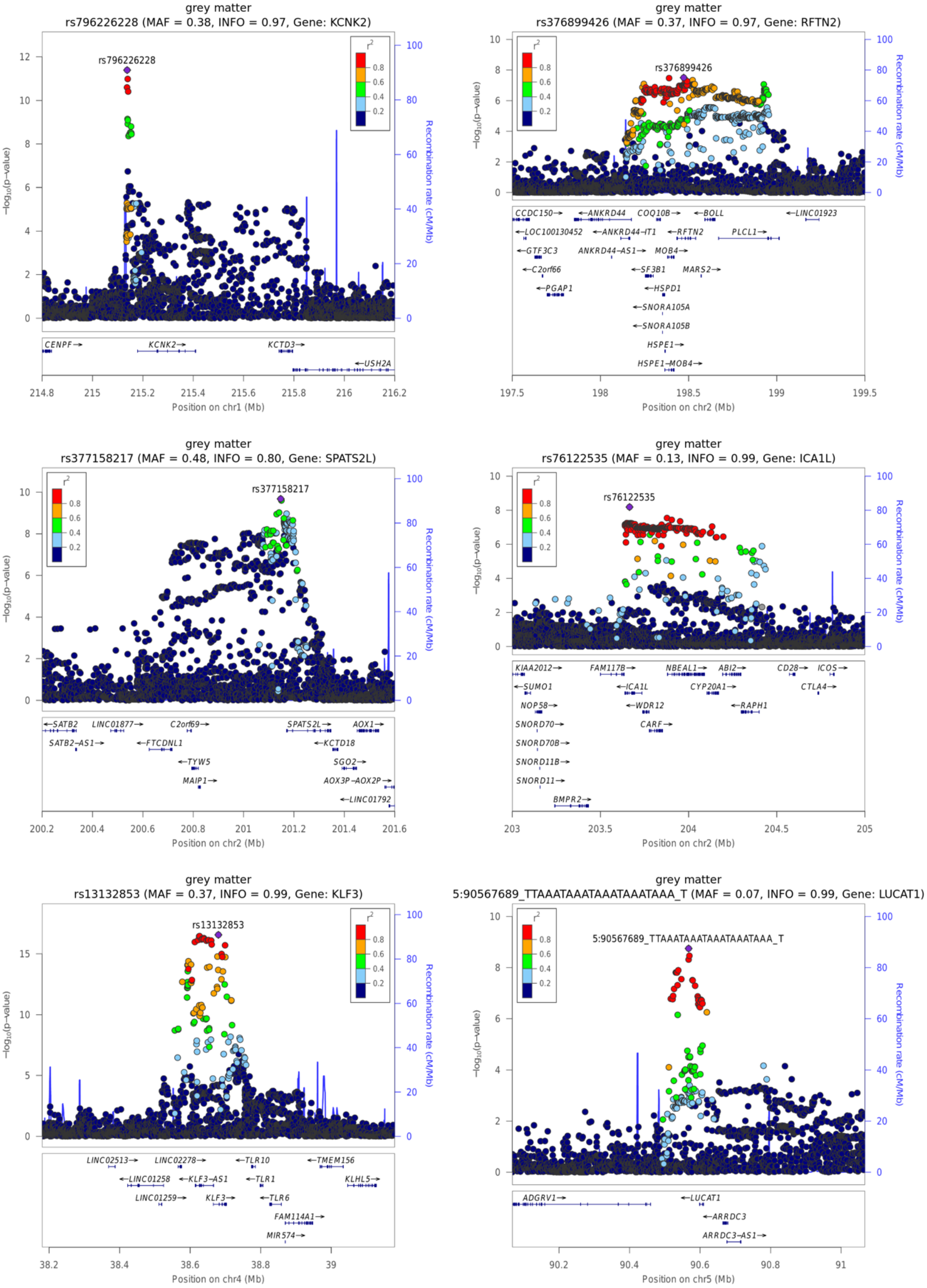


**Fig. A5** Regional association plots for index variations 1-6 from the genome-wide association analysis of grey matter brain age gap in N = 32,634 white-British ancestry individuals. Regional association plots were created using Locuszoom Standalone v1.4. SNP positions (dbSNP build 151) and refFlat gene locations (2020-08-17) are based on human genome build hg19 and were accessed via UCSC Genome Browser. Recombination rates were derived from HapMap phase II build GRCh37 (2011-01-19). MAF: Minor allele frequency, INFO: Imputation Information Score, $r^2$: linkage disequilibrium between index variation and other variation in locus.

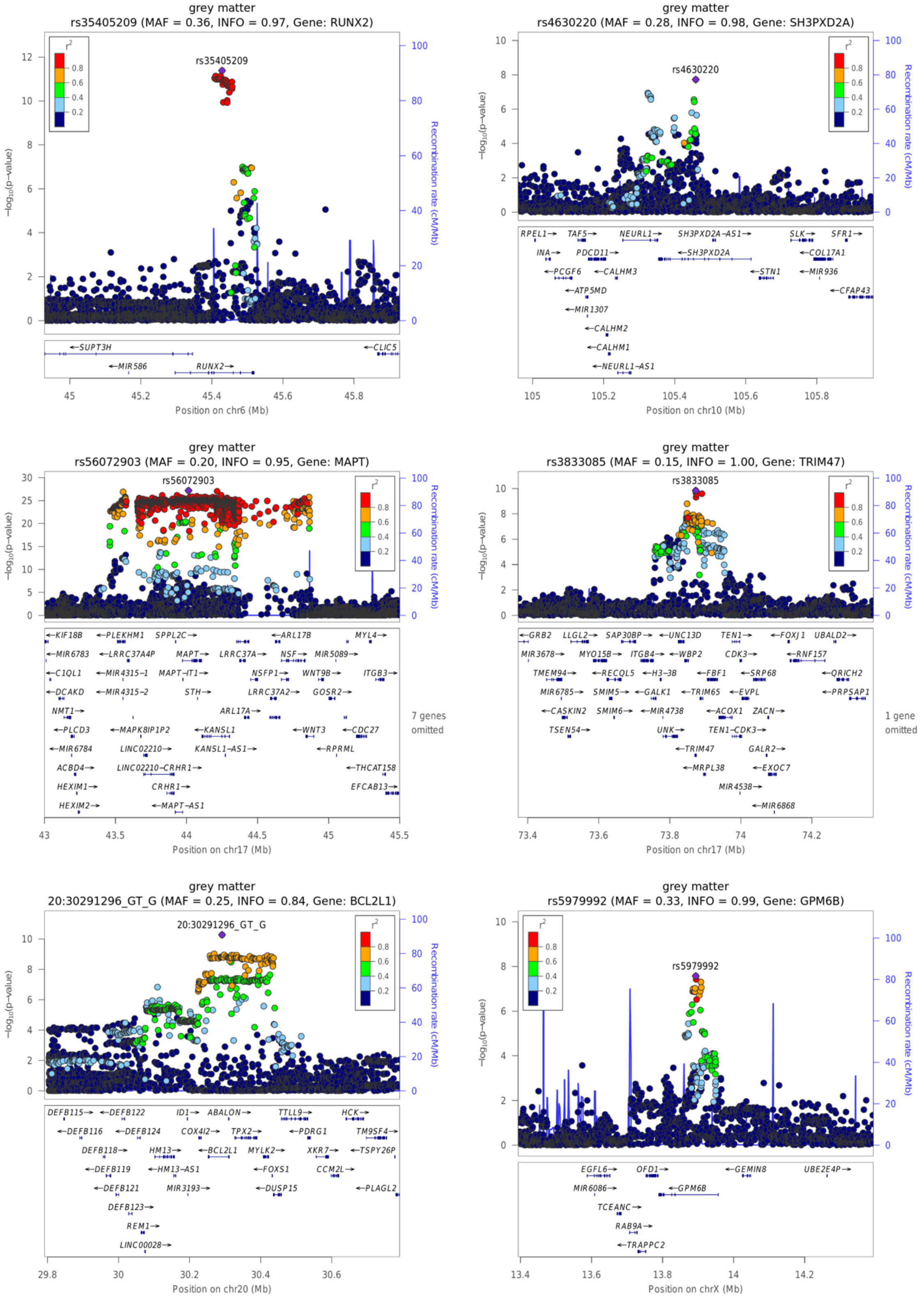


**Fig. A6** Regional association plots for index variations 7-12 from the genome-wide association analysis of grey matter brain age gap in N = 32,634 white-British ancestry individuals. Regional association plots were created using Locuszoom Standalone v1.4. SNP positions (dbSNP build 151) and refFlat gene locations (2020-08-17) are based on human genome build hg19 and were accessed via UCSC Genome Browser. Recombination rates were derived from HapMap phase II build GRCh37 (2011-01-19). MAF: Minor allele frequency, INFO: Imputation Information Score, $r^2$: linkage disequilibrium between index variation and other variation in locus.

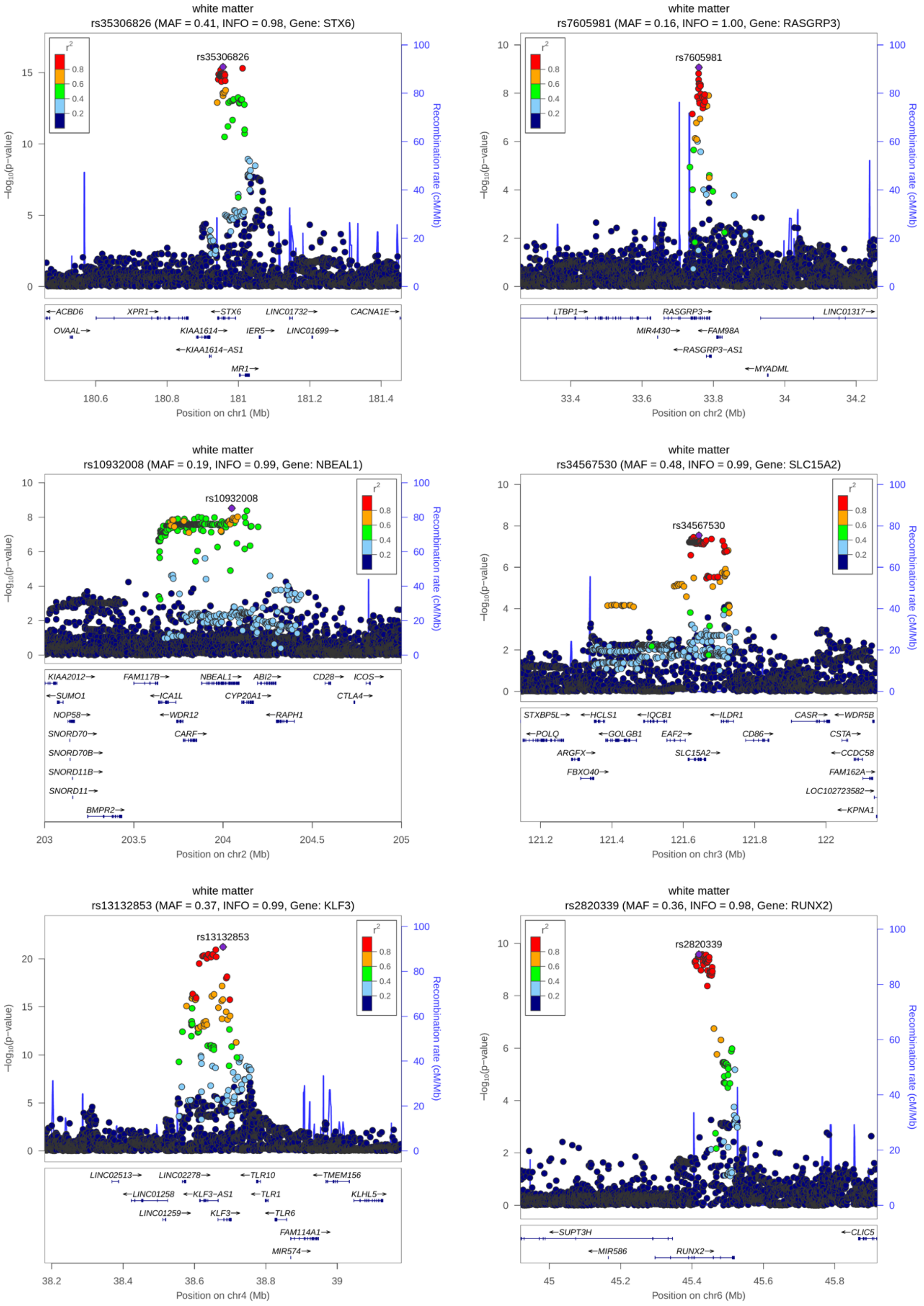


**Fig. A7** Regional association plots for index variations 1-6 from the genome-wide association analysis of white matter brain age gap in N = 32,634 white-British ancestry individuals. Regional association plots were created using Locuszoom Standalone v1.4. SNP positions (dbSNP build 151) and refFlat gene locations (2020-08-17) are based on human genome build hg19 and were accessed via UCSC Genome Browser. Recombination rates were derived from HapMap phase II build GRCh37 (2011-01-19). MAF: Minor allele frequency, INFO: Imputation Information Score, $r^2$: linkage disequilibrium between index variation and other variation in locus.

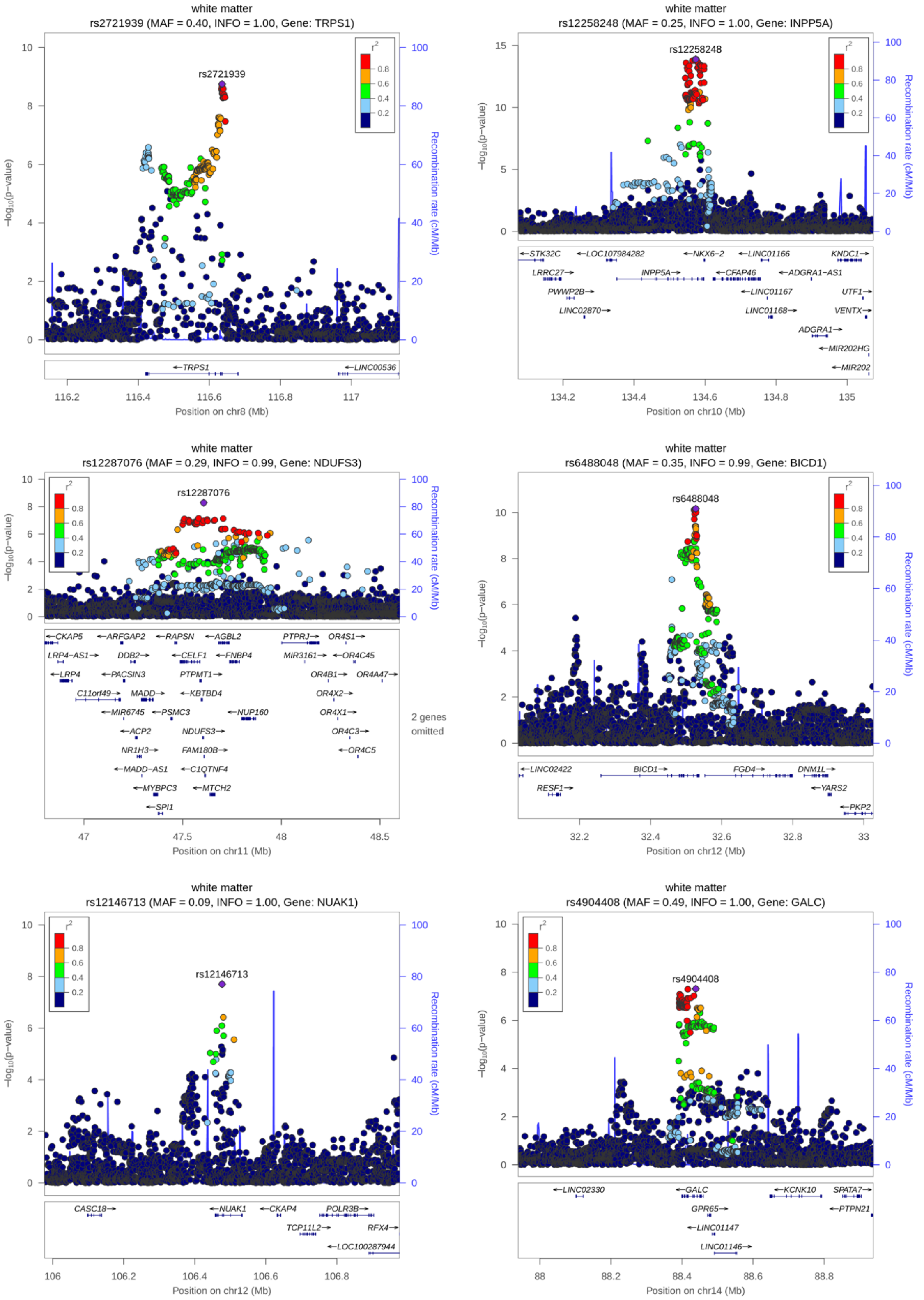


**Fig. A8** Regional association plots for index variations 7-12 from the genome-wide association analysis of white matter brain age gap in N = 32,634 white-British ancestry individuals. Regional association plots were created using Locuszoom Standalone v1.4. SNP positions (dbSNP build 151) and refFlat gene locations (2020-08-17) are based on human genome build hg19 and were accessed via UCSC Genome Browser. Recombination rates were derived from HapMap phase II build GRCh37 (2011-01-19). MAF: Minor allele frequency, INFO: Imputation Information Score, $r^2$: linkage disequilibrium between index variation and other variation in locus.

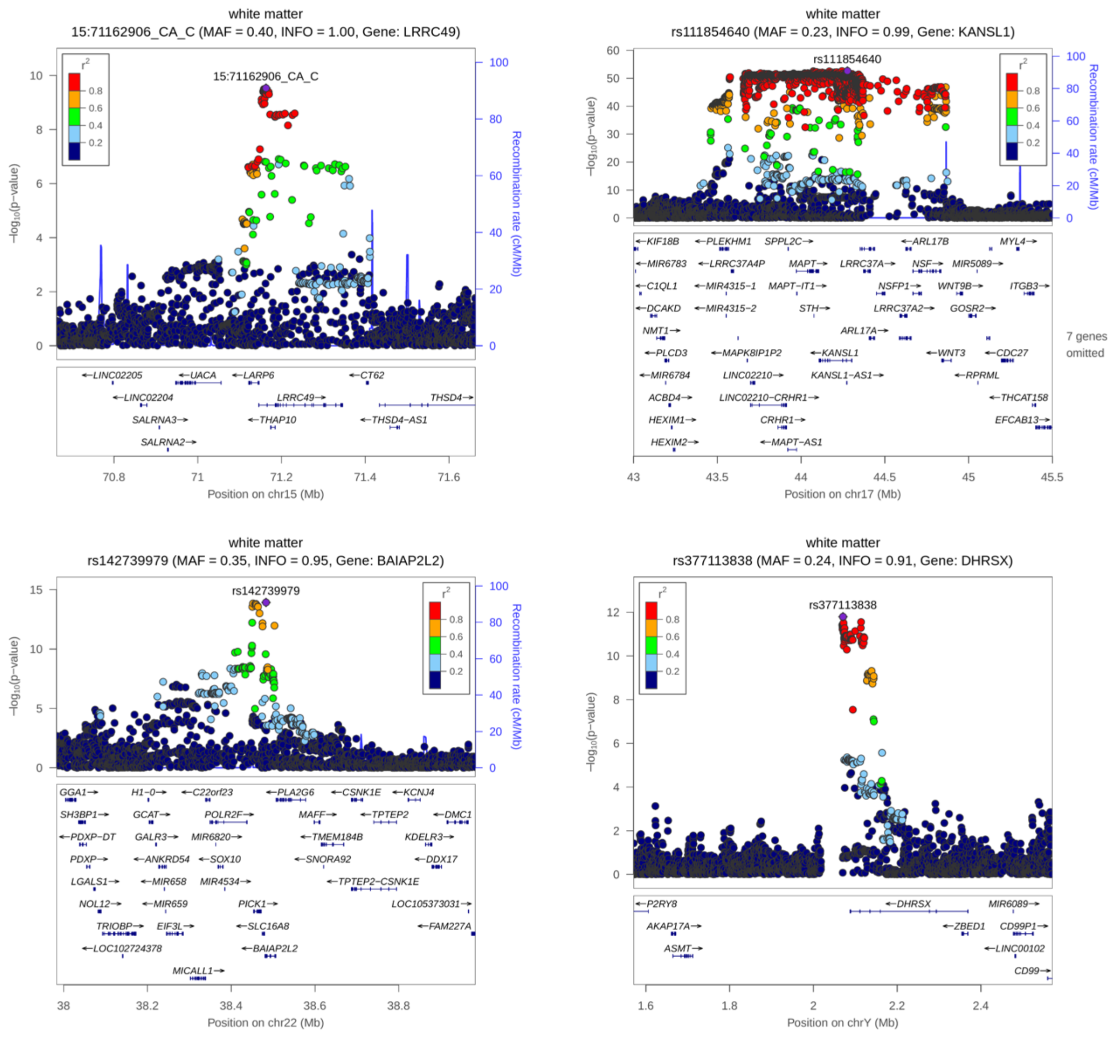


**Fig. A9** Regional association plots for index variations 13-16 from the genome-wide association analysis of white matter brain age gap in N = 32,634 white-British ancestry individuals. Regional association plots were created using Locuszoom Standalone v1.4. SNP positions (dbSNP build 151) and refFlat gene locations (2020-08-17) are based on human genome build hg19 and were accessed via UCSC Genome Browser. Recombination rates were derived from HapMap phase II build GRCh37 (2011-01-19). MAF: Minor allele frequency, INFO: Imputation Information Score, $r^2$: linkage disequilibrium between index variation and other variation in locus.

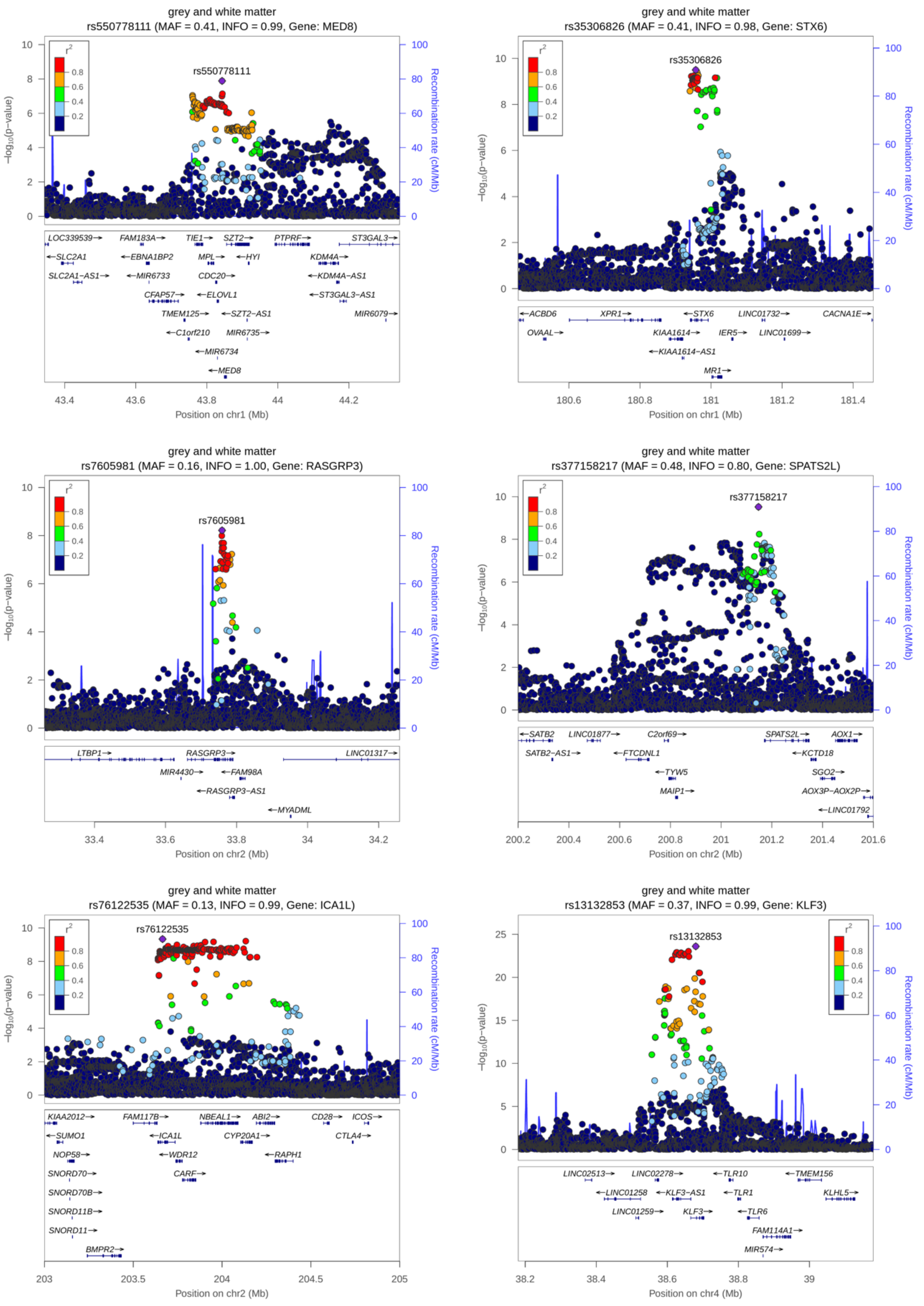


**Fig. A10** Regional association plots for index variations 1-6 from the genome-wide association analysis of the combined grey and white matter brain age gap estimates in N = 32,634 white-British ancestry individuals. Regional association plots were created using Locuszoom Standalone v1.4. SNP positions (dbSNP build 151) and refFlat gene locations (2020-08-17) are based on human genome build hg19 and were accessed via UCSC Genome Browser. Recombination rates were derived from HapMap phase II build GRCh37 (2011-01-19). MAF: Minor allele frequency, INFO: Imputation Information Score, $r^2$: linkage disequilibrium between index variation and other variation in locus.

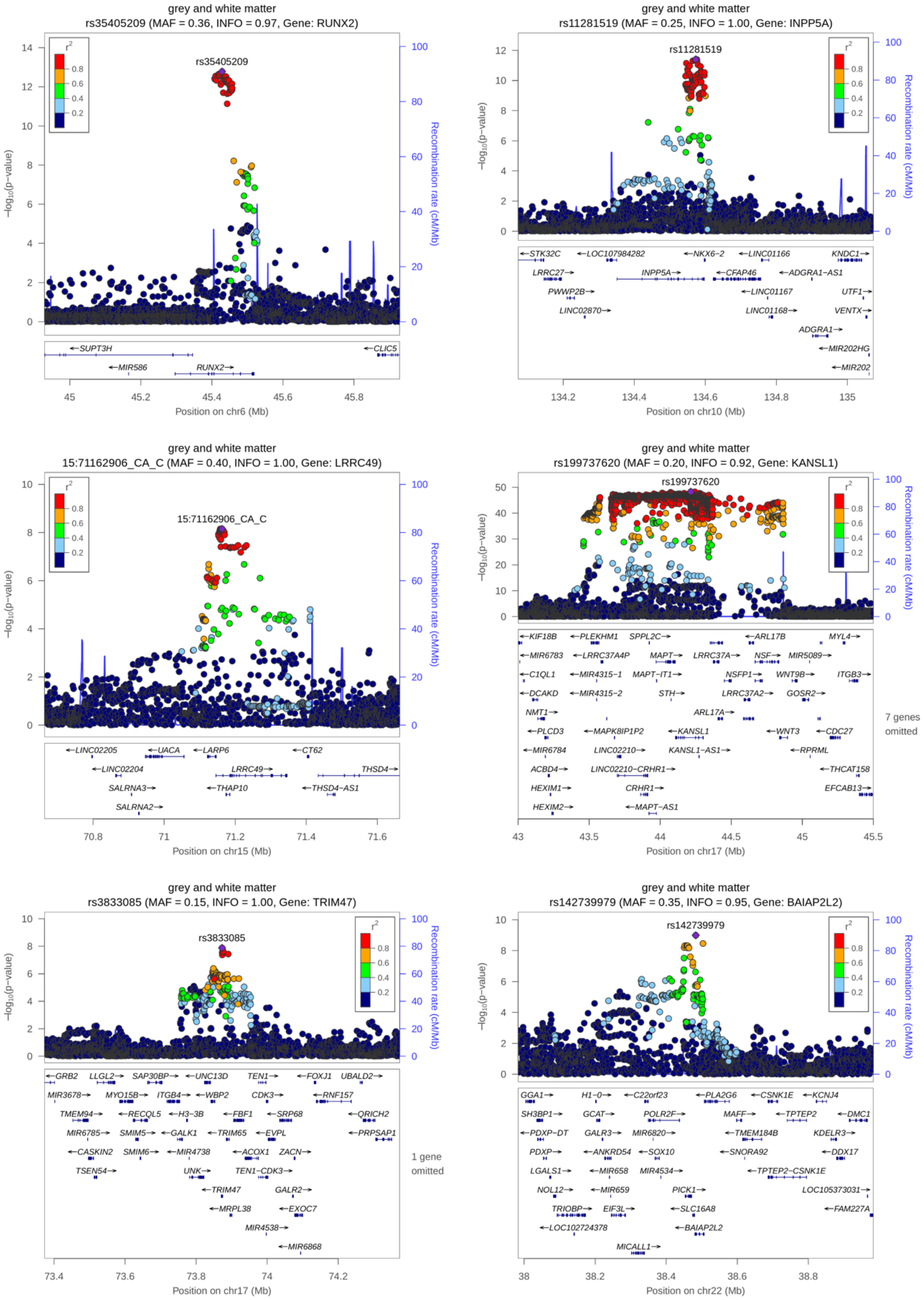


**Fig. A11** Regional association plots for index variations 7-12 from the genome-wide association analysis of the combined grey and white matter brain age gap estimates in N = 32,634 white-British ancestry individuals. Regional association plots were created using Locuszoom Standalone v1.4. SNP positions (dbSNP build 151) and refFlat gene locations (2020-08-17) are based on human genome build hg19 and were accessed via UCSC Genome Browser. Recombination rates were derived from HapMap phase II build GRCh37 (2011-01-19). MAF: Minor allele frequency, INFO: Imputation Information Score, $r^2$: linkage disequilibrium between index variation and other variation in locus.

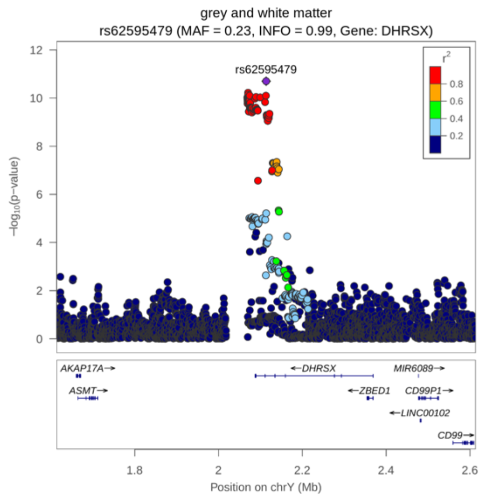


**Fig. A12** Regional association plot for index variation 13 from the genome-wide association analysis of the combined grey and white matter brain age gap estimates in N = 32,634 white-British ancestry individuals. Regional association plots were created using Locuszoom Standalone v1.4. SNP positions (dbSNP build 151) and refFlat gene locations (2020-08-17) are based on human genome build hg19 and were accessed via UCSC Genome Browser. Recombination rates were derived from HapMap phase II build GRCh37 (2011-01-19). MAF: Minor allele frequency, INFO: Imputation Information Score, $r^2$: linkage disequilibrium between index variation and other variation in locus.

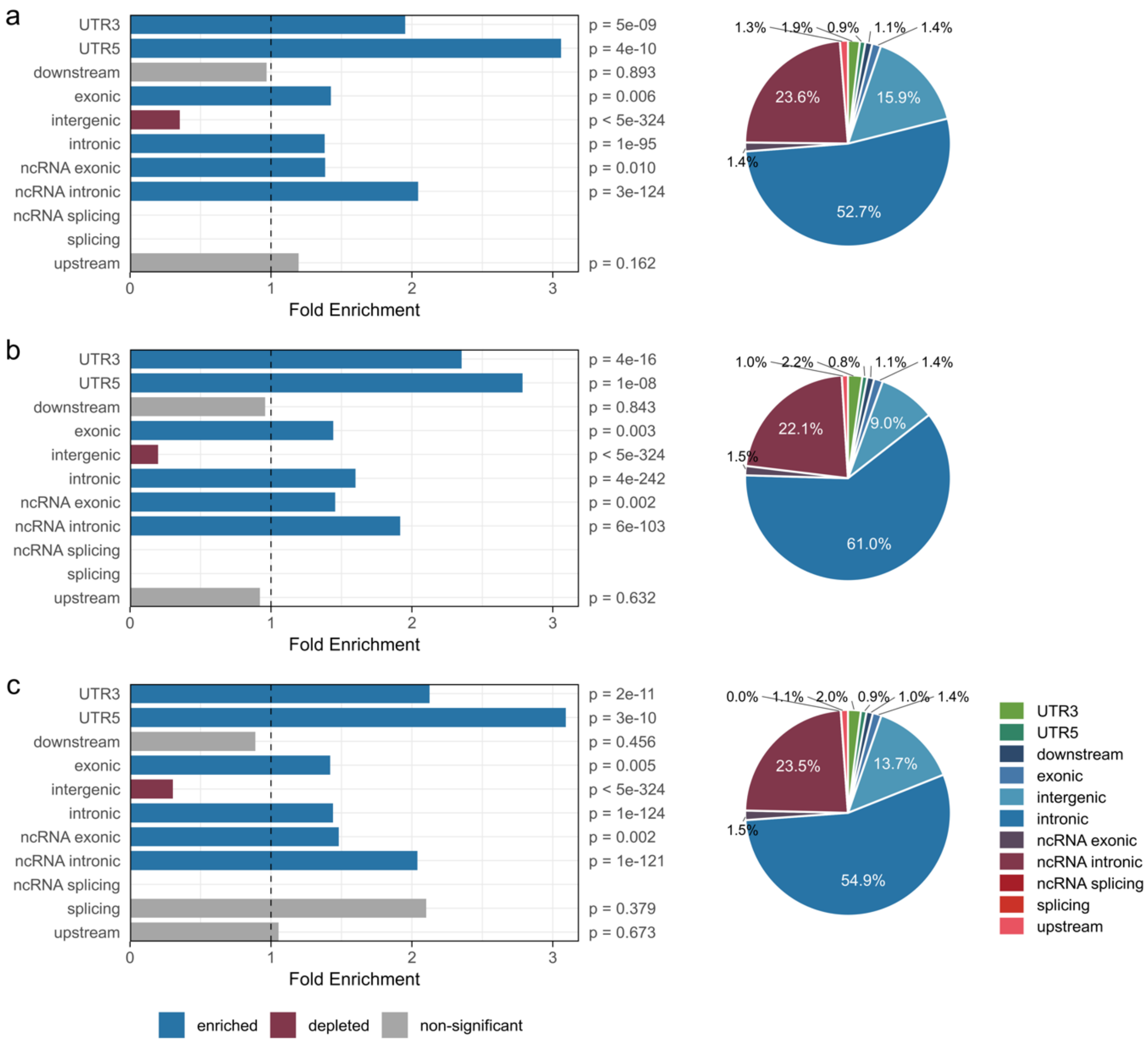


**Fig. A13** Results of the ANNOVAR enrichment test of functional consequences of discovered variations. All 'candidate-SNPs' (see methods for definition) identified by FUMA in genome-wide significant loci have been considered for the ANNOVAR enrichment test. Bar diagrams show the Fold Enrichment (*FE*), i.e., the ratio of the observed proportion vs. the expected proportion of candidate-SNPs annotated with the respective functional consequence. *FE* values higher than 1 indicate an enrichment, whereas *FE* values lower than 1 indicate a depletion of the respective functional consequence. Pie charts show the distribution of variations annotated with the respective functional consequences. **(a)** grey matter brain age gap (4,945 candidate-SNPs), **(b)** white matter brain age gap (5,177 candidate-SNPs), and **(c)** combined grey and white matter brain age gap (4,890 candidate-SNPs).

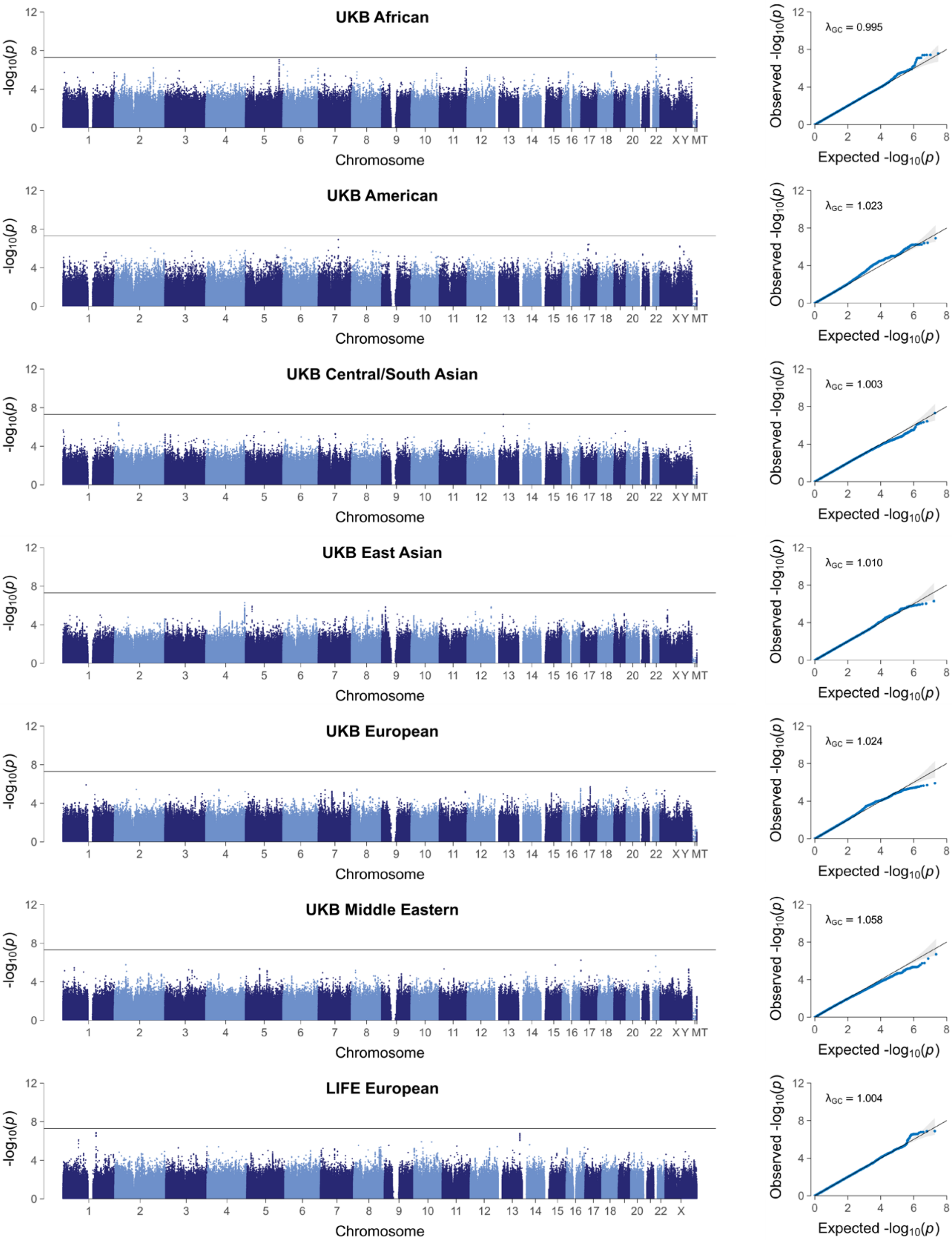


**Fig. A14** Manhattan plots (left) and quantile-quantile plots (right) showing the results of the genome-wide association analyses for grey matter brain age gap in the seven replication samples: UKB African ancestry ($n = 217$), UKB Admixed American ancestry ($n = 60$); UKB Central/South Asian ancestry ($n = 409$), UKB East Asian ancestry ($n = 192$), UKB European ancestry ($n = 4{,}486$); UKB Middle Eastern ancestry ($n = 62$); LIFE-Adult European ancestry ($n = 1{,}833$). Manhattan plots show the $p$-values (-$\log_{10}$ scale) of the tested genetic variations on the y-axis and base-pair positions along the chromosomes on the x-axis. The solid horizontal line indicates the threshold of genome-wide significance ($p$ = 5E-8). Pseudoautosomal variations have been added to chromosome 'X'. Quantile-quantile plots show the observed $p$-values from the association analysis vs. the expected $p$-values under the null hypothesis of no effect (-$\log_{10}$ scale).

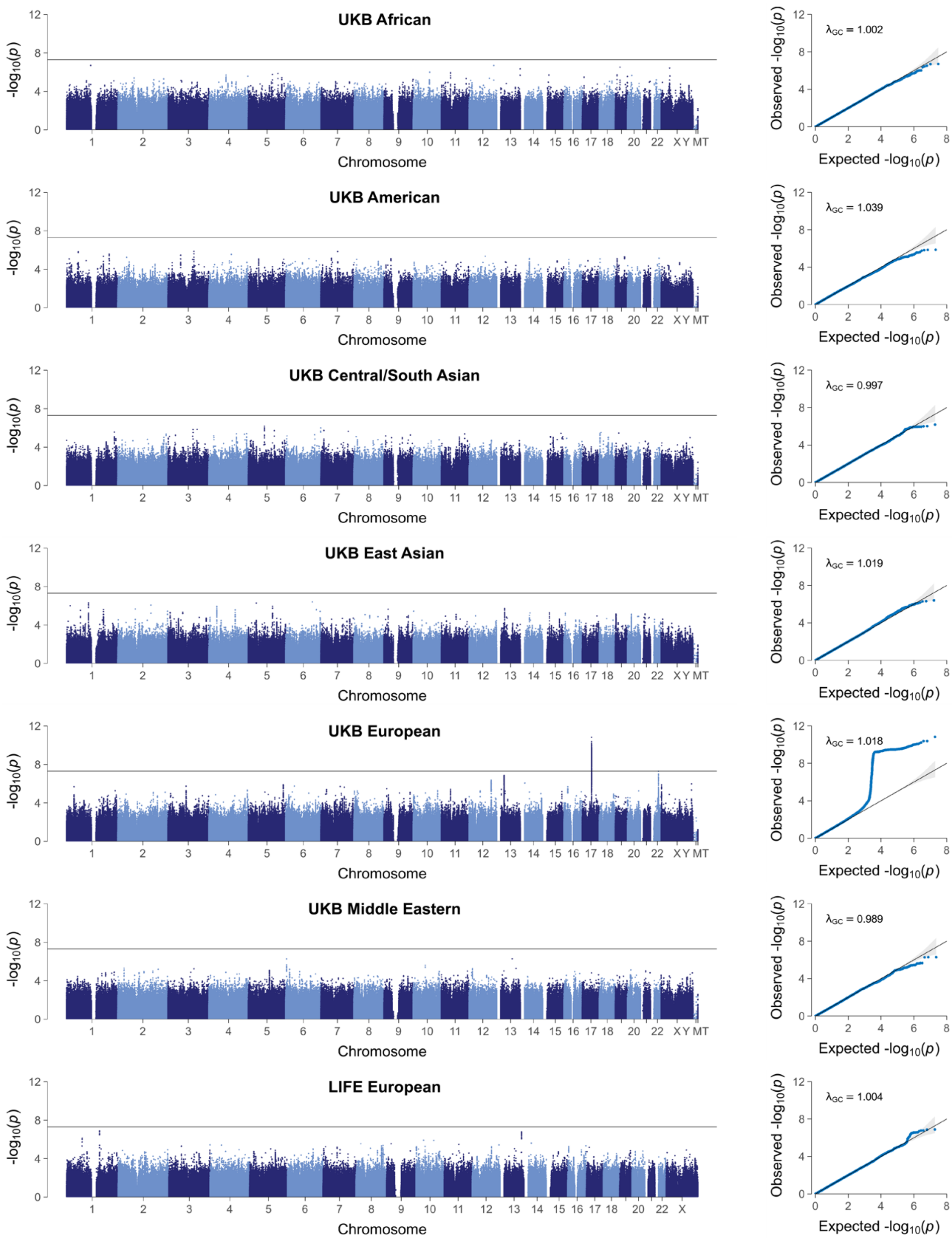


**Fig. A15** Manhattan plots (left) and quantile-quantile plots (right) showing the results of the genome-wide association analyses for white matter brain age gap in the seven replication samples: UKB African ancestry ($n = 217$), UKB Admixed American ancestry ($n = 60$); UKB Central/South Asian ancestry ($n = 409$), UKB East Asian ancestry ($n = 192$), UKB European ancestry ($n = 4{,}486$); UKB Middle Eastern ancestry ($n = 62$); LIFE-Adult European ancestry ($n = 1{,}883$). Manhattan plots show the $p$-values (-$\log_{10}$ scale) of the tested genetic variations on the y-axis and base-pair positions along the chromosomes on the x-axis. The solid horizontal line indicates the threshold of genome-wide significance ($p$ = 5E-8). Pseudoautosomal variations have been added to chromosome 'X'. Quantile-quantile plots show the observed $p$-values from the association analysis vs. the expected $p$-values under the null hypothesis of no effect (-$\log_{10}$ scale).

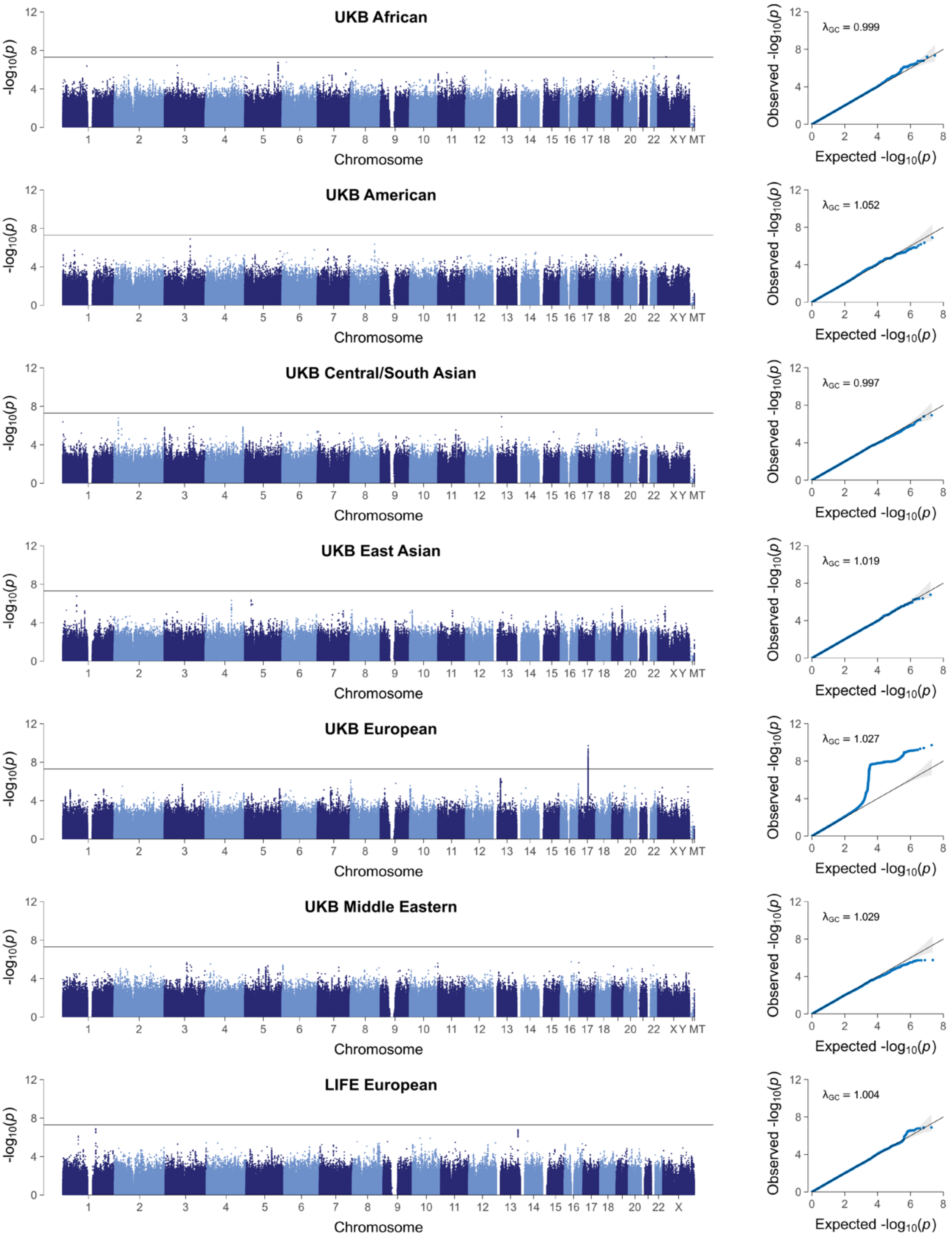


**Fig. A16** Manhattan plots (left) and quantile-quantile plots (right) showing the results of the genome-wide association analyses for combined grey and white matter brain age gap in the seven replication samples: UKB African ancestry ($n = 217$), UKB Admixed American ancestry ($n = 60$); UKB Central/South Asian ancestry ($n = 409$), UKB East Asian ancestry ($n = 192$), UKB European ancestry ($n = 4{,}486$); UKB Middle Eastern ancestry ($n = 62$); LIFE-Adult European ancestry ($n = 1{,}883$). Manhattan plots show the $p$-values (-log$_{10}$ scale) of the tested genetic variations on the y-axis and base-pair positions along the chromosomes on the x-axis. The solid horizontal line indicates the threshold of genome-wide significance ($p$ = 5E-8). Pseudoautosomal variations have been added to chromosome ‘X’. Quantile-quantile plots show the observed $p$-values from the association analysis vs. the expected $p$-values under the null hypothesis of no effect (-log$_{10}$ scale).

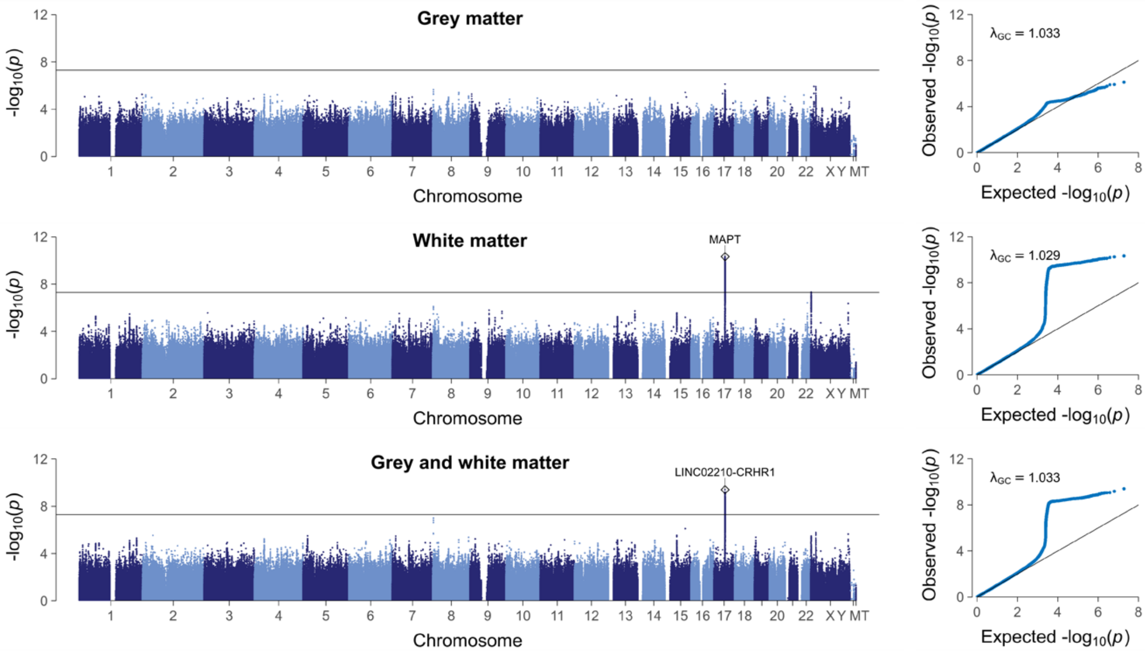


**Fig. A17** Manhattan plots (left) and quantile-quantile plots (right) showing the results of the cross-ancestry replication genome-wide association meta-analyses (GWAMAs) for the three brain age gap traits. Replication GWAMA results combine individual GWAS results from up to seven samples: UKB African ancestry ($n = 217$), UKB Admixed American ancestry ($n = 60$); UKB Central/South Asian ancestry ($n = 409$), UKB East Asian ancestry ($n = 192$), UKB European ancestry ($n = 4{,}486$), UKB Middle Eastern ancestry ($n = 62$), LIFE-Adult European ancestry ($n = 1{,}883$). Manhattan plots show the $p$-values (-log$_{10}$ scale) of the tested genetic variations on the y-axis and base-pair positions along the chromosomes on the x-axis. The solid horizontal line indicates the threshold of genome-wide significance ($p$ = 5E-8). Pseudoautosomal variations have been added to chromosome 'X'. Quantile-quantile plots show the observed $p$-values from the association analysis vs. the expected $p$-values under the null hypothesis of no effect (-log$_{10}$ scale).

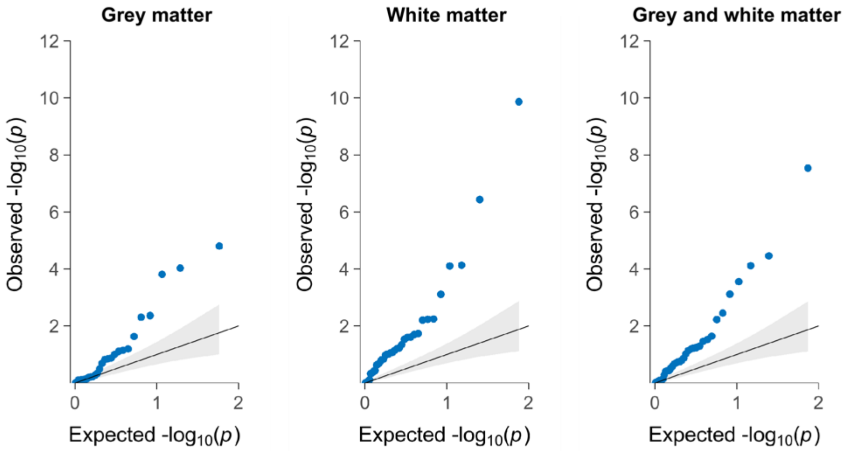


**Fig. A18** Quantile-quantile plots showing the replication results for independent variants with discovery $p <$ 1.0e-06. Quantile-quantile plots show the observed vs. expected $p$-values (two-tailed) from the cross-ancestry replication meta-analysis in up to $N$ = 7,259 individuals. Blue dots reflect the observed p-values sorted from largest to smallest and plotted against the expected p-values under the null hypothesis of no effect (-log$_{10}$ scale). The solid diagonal line reflects the mean expected p-values. The lower and upper bound of the grey shaded area represent the 5th and 95th percentile of the expected p-values. The quantile-quantile plots show an excess of low $p$-values observed in replication analyses, suggesting stronger evidence than expected under the null.

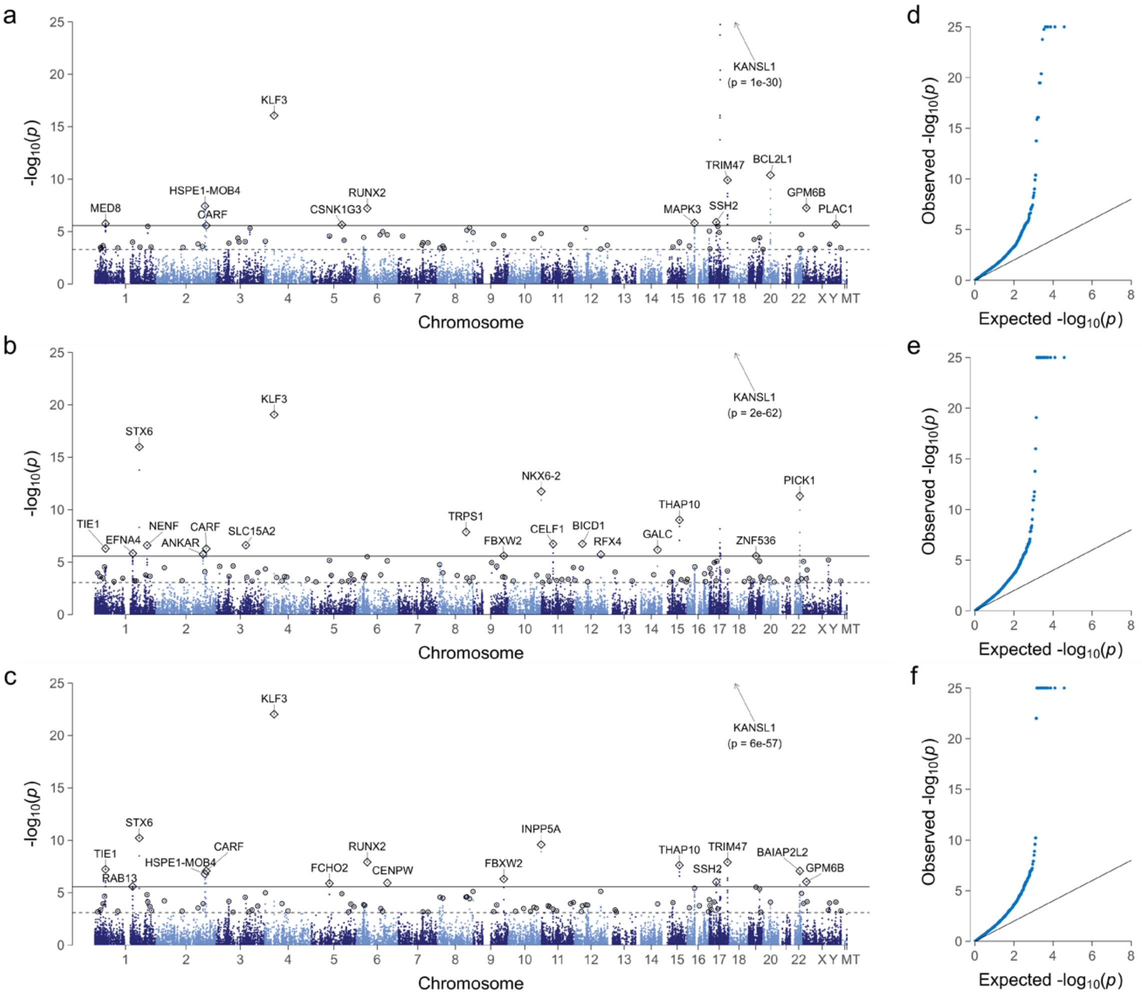


**Fig. A19** Manhattan plots (**a-c**) and quantile-quantile plots (**d-f**) showing the results of the gene-based association analyses for the three brain age gap traits ($N = 32{,}634$ UK Biobank individuals). Manhattan plots show the $p$-values (-$\log_{10}$ scale) of the tested genes on the y-axis and base-pair positions (gene start coordinates) along the chromosomes on the x-axis. In total, 18,634 protein-coding genes (RefSeq assembly GRCh37.p13, 09-05-2019) have been included. The solid horizontal line reflects the Bonferroni-corrected level of significance. The dashed horizontal line reflects the FDR-corrected level of significance. Diamonds and circles highlight the index gene in each genomic locus (diamond: Bonferroni-significant index gene; circles: FDR-significant index gene). Index genes reaching the Bonferroni-corrected level of significance have been annotated with respective gene symbols. Genes in a physical distance of 10 Mbp were regarded to lie in the same genomic locus and are thus represented by the same index gene. Quantile-quantile plots show the observed $p$-values from the association analysis vs. the expected $p$-values under the null hypothesis of no effect (-$\log_{10}$ scale). For illustrative reasons, the y-axis has been truncated at $p = $ 1E-25. **a,d** grey matter brain age gap; **b,e** white matter brain age gap; **c,f** combined grey and white matter brain age gap.

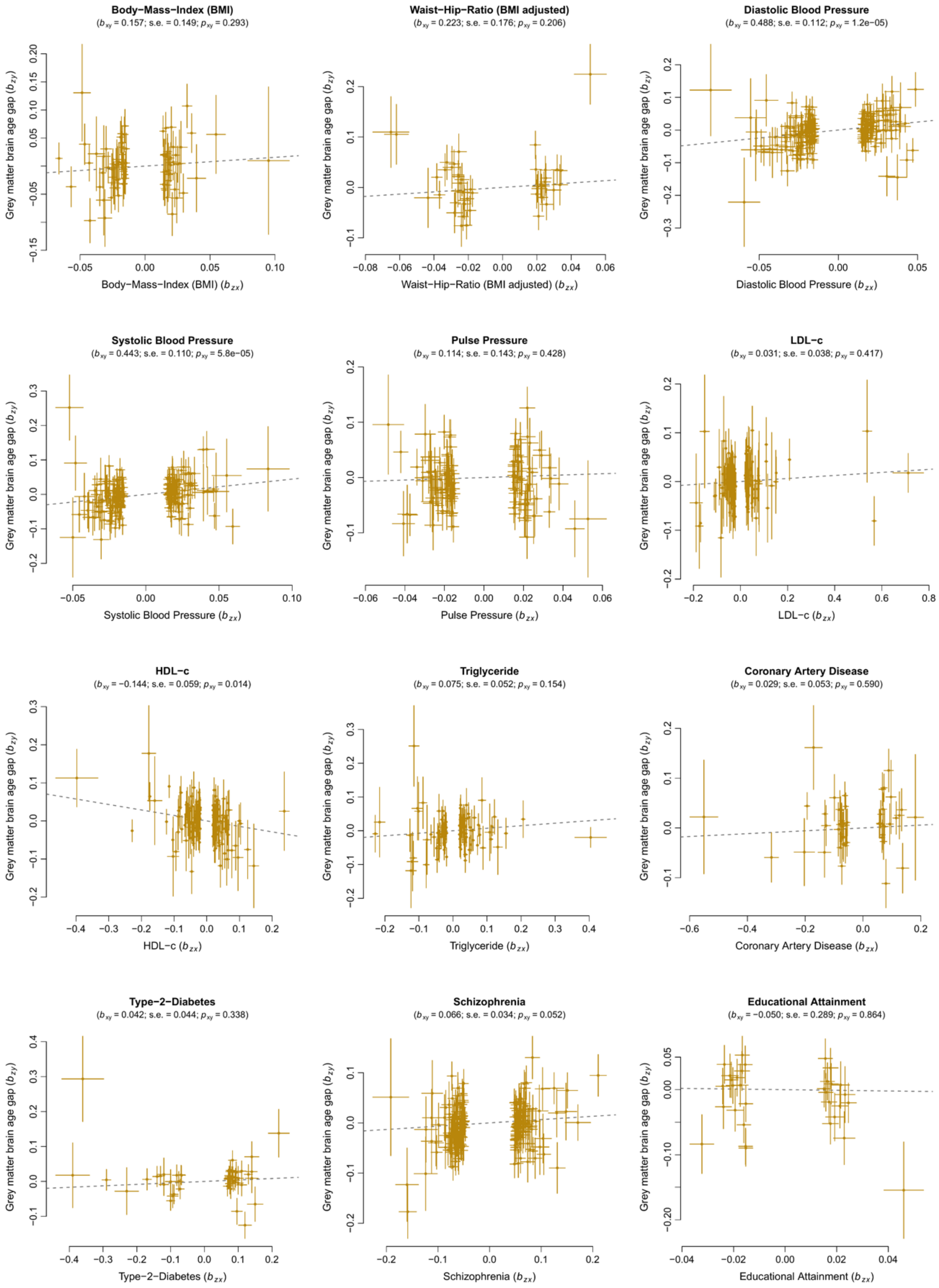


**Fig. A20** Results of the generalized summary-data-based Mendelian Randomization (GSMR) analyses for grey matter brain age gap and 11 modifiable risk factors. Each plot shows multiple genetic variants serving as instruments to test for causality between an exposure (risk factor) and an outcome variable (brain age gap). Under a causal model, variant effects on the outcome ($b_{\mathrm{zy}}$; y-axis) are expected to be linearly proportional to the variant effects on the exposure variable ($b_{\mathrm{zx}}$; x-axis). The ratio between $b_{\mathrm{zy}}$ and $b_{\mathrm{zx}}$ provides an estimate of the mediation effect of x on y ($b_{\mathrm{xy}}$). Variants with potential horizontal pleiotropic effects were identified using the HEIDI-outlier method and were removed in advance. s.e. standard error of the mediation effect; $p_{\mathrm{xy}}$ p-value of the mediation effect

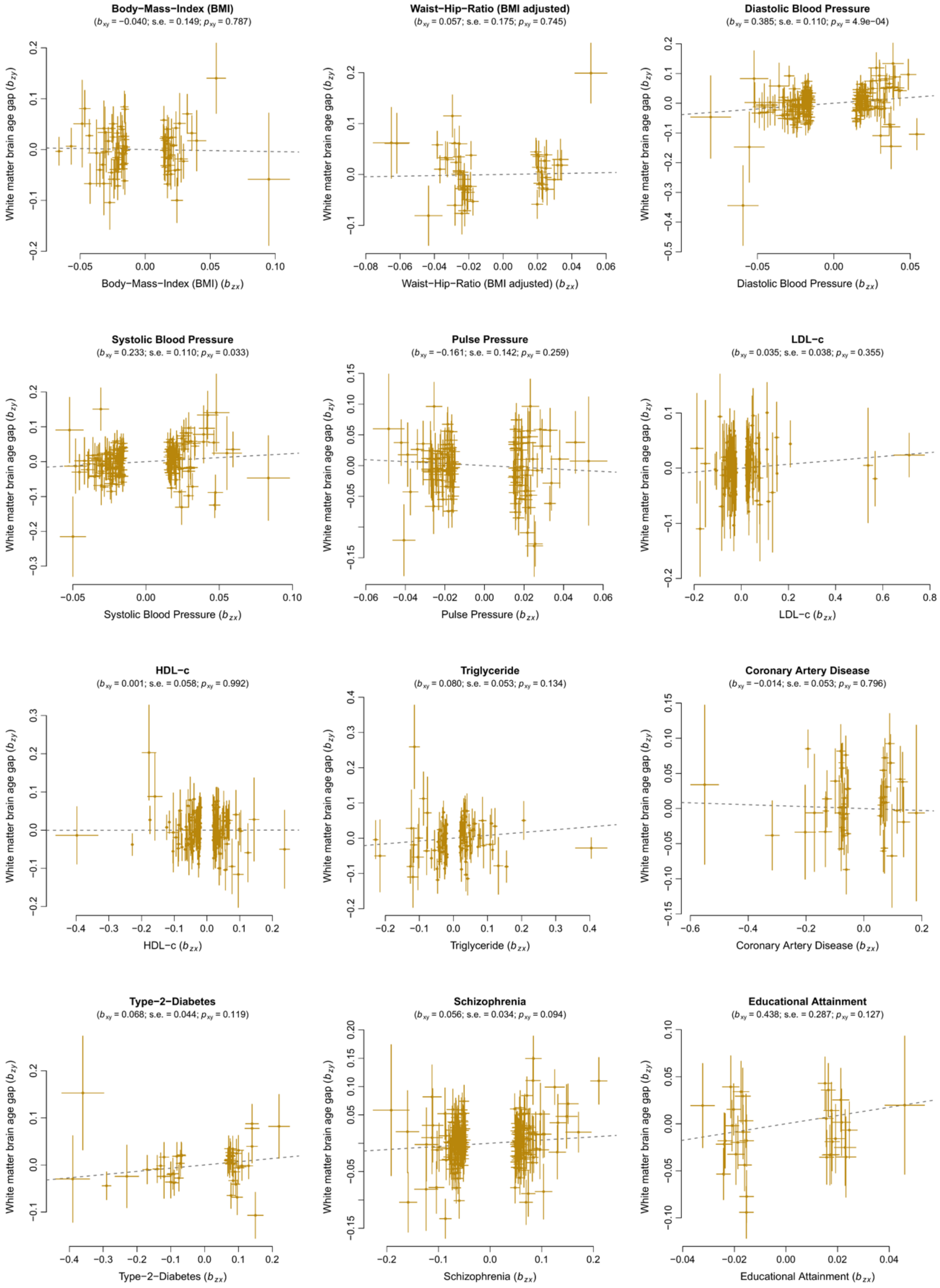


**Fig. A21** Results of the generalized summary-data-based Mendelian Randomization (GSMR) analyses for white matter brain age gap and 11 modifiable risk factors. Each plot shows multiple genetic variants serving as instruments to test for causality between an exposure (risk factor) and an outcome variable (brain age gap). Under a causal model, variant effects on the outcome ($b_{zy}$; y-axis) are expected to be linearly proportional to the variant effects on the exposure variable ($b_{zx}$; x-axis). The ratio between $b_{zy}$ and $b_{zx}$ provides an estimate of the mediation effect of x on y ($b_{xy}$). Variants with potential horizontal pleiotropic effects were identified using the HEIDI-outlier method and were removed in advance. s.e. standard error of the mediation effect; $p_{xy}$ p-value of the mediation effect

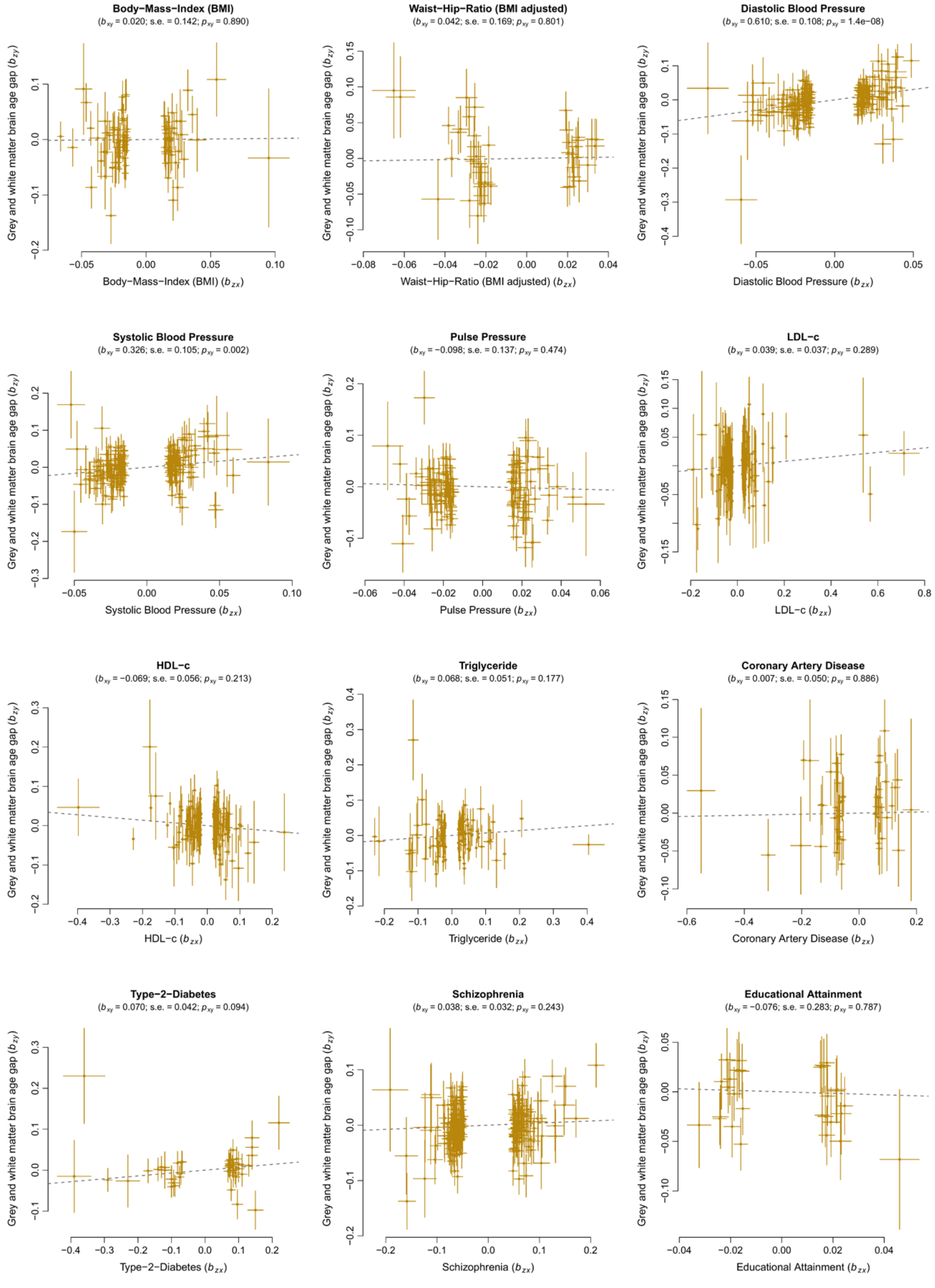


**Fig. A22** Results of the generalized summary-data-based Mendelian Randomization (GSMR) analyses for combined grey and white matter brain age gap and 11 modifiable risk factors. Each plot shows multiple genetic variants serving as instruments to test for causality between an exposure (risk factor) and an outcome variable (brain age gap). Under a causal model, variant effects on the outcome ($b_{\mathrm{zy}}$; y-axis) are expected to be linearly proportional to the variant effects on the exposure variable ($b_{\mathrm{zx}}$; x-axis). The ratio between $b_{\mathrm{zy}}$ and $b_{\mathrm{zx}}$ provides an estimate of the mediation effect of x on y ($b_{\mathrm{xy}}$). Variants with potential horizontal pleiotropic effects were identified using the HEIDI-outlier method and were removed in advance. s.e. standard error of the mediation effect; $p_{\mathrm{xy}}$ p-value of the mediation effect

### References

McGraw, K. O., and Wong, S. P. (1996). Forming inferences about some intraclass correlation coefficients. *Psychol. Methods* 1, 30–46. doi:10.1037/1082-989X.1.1.30.
